# EXERCISE IN WOMEN UNDERGOING BARIATRIC SURGERY: IMPACT ON THE OVARIAN FUNCTION AND CARDIOMETABOLIC RISK FACTORS – THE EMOVAR STUDY

**DOI:** 10.1101/2025.11.16.25340335

**Authors:** Andrés Baena-Raya, Sonia Martínez-Forte, Elena Martínez-Rosales, Manuel Ferrer-Márquez, Laura López-Sánchez, Alba Hernández-Martínez, Alba Esteban-Simón, David Ruiz-González, Pablo Soriano-Maldonado, Lorena Carmona-Rodríguez, Ana del Mar Salmerón, Ana Cristina Abreu, Jesús Aceituno-Cubero, Manuel A. Rodríguez-Pérez, Carlos Gómez-Navarro, Ignacio Fernández-de-las-Nieves, Enrique García-Artero, Borja Martínez-Téllez, Ana M. Fernández-Alonso, Alberto Soriano-Maldonado

## Abstract

**Backgroud:** This study aimed to assess the effects of a 16-week supervised exercise intervention on ovarian function in women undergoing bariatric surgery (BS); and to examine potential mechanisms associated with the changes in ovarian function.

**Materials and Methods:** A randomized, two-arm parallel-group trial was conducted from October 2019 to September 2022. Participants were reproductive-aged women with severe obesity (BMI ≥40 kg/m^2^ or BMI ≥35 with comorbidities) recruited from the BS services from two hospitals. Participants were randomly assigned to BS+usual care (*n* = 25) or BS+exercise (*n* = 21), consisting of 16 weeks of three-weekly supervised exercise sessions. Outcomes were assessed before surgery, at week 16, and 1-year. The primary outcome was the change in sex-hormone binging globulin (SHBG). Secondary outcomes were related to ovarian function (obtained from both serum and transvaginal ultrasound), weight loss, body composition, fitness, inflammation, cardiometabolic and nuclear magnetic resonance-derived metabolomic profiles.

**Results:** A total of 42 participants (91%; 18 in BS+EX; 24 in BS+usual care) were included in the primary analyses. There were no between-group differences at week 16. At 1-year, the exercise group increased serum SHBG levels (+36.3 nmol/L; 95%CI 2.3 to 70.2; *p=*0.037), oocyte count (+3.1 follicles; 95%CI 0.9 to 5.3; *p=*0.007), and reduced the uterine artery mean pulsatility index (UtA-PI) (−1.3; 95%CI −2.1 to −0.5; *p=*0.003) compared to usual care, despite comparable weight loss and changes in secondary/exploratory outcomes. The 1-year changes in metabolomic profile predicted 97% of the increase in SHBG in the exercise group. Interestingly, the decrease in serum amino acid levels was associated with increased SHBG levels at 1-year, only in the exercise group. Sensitivity analyses corroborated the results.

**Conclusion:** The EMOVAR trial suggests that a 16-week supervised exercise program improves relevant markers of ovarian function, such as SHBG, oocyte count, and UtA-PI, at 1-year compared to usual care.

## INTRODUCTION

Severe obesity disrupts the ovarian function and causes ovulatory disorders and infertility in about 48.5 million women aged 20-44 worldwide ^[1]^. Ovulatory disorders are the primary clinical manifestation in women with obesity, mainly due to neuroendocrine dysfunction within the hypothalamic-pituitary-ovarian axis ^[2,3]^. Insulin resistance in women with obesity increases peripheral conversion of androgens to estrogens and decreases hepatic synthesis of sex hormone-binding globulin (SHBG) ^[4,5]^. SHBG binds free testosterone and estrogen to regulate sex steroid levels. Lower SHBG levels lead to higher free estrogen and testosterone, negatively affecting follicle-stimulating hormone (FSH) secretion and potentially inhibiting ovulation ^[6]^. Similarly, excess adiposity increases lipotoxicity and markers of systemic inflammation such as C-reactive protein (CRP), leptin, or tumor necrosis factor-alpha (TNF-α) ^[7]^. These metabolic modifications may disrupt oocyte maturation and follicular development, leading to menstrual irregularities and ovulatory dysfunction ^[2,7]^.

Bariatric surgery (BS) is the most effective intervention for achieving significant weight loss ^[8]^, resulting in reduced cardiovascular risk factors ^[9]^, improved metabolomic profile ^[10]^ and better quality of life ^[11]^. Evidence supports that BS might also improve ovarian function by increasing SHBG ^[12]^ and reducing total serum testosterone ^[13]^ levels at 6 and 12 months post-surgery. A 5-year retrospective study ^[14]^ of 932 women who underwent BS also showed lower testosterone levels and reduced menstrual irregularities. Nevertheless, these benefits might be reversed due to recurrent weight gain, which affects up to 50% of BS patients ^[15]^. In this context, exercise is suggested as a complementary therapy. In women with obesity, exercise alone enhances the ovarian function and regulates menstrual cycles ^[16]^ by decreasing the free androgen index (FAI) ^[17]^ and increasing SHBG ^[18]^.

Ruiz-Gonzalez et al. ^[19]^ revealed that, in women with overweight or obesity, adding exercise within a holistic approach that includes pharmacological and nutritional strategies, regulates the reproductive hormonal profile and restores ovulation rates. Exercise following BS further benefits cardiorrespiratory fitness ^[20]^ and cardiometabolic risk factors ^[21,22]^ compared to usual care, even without weight loss or body composition changes ^[23]^. Thus, we hypothesize that exercise could enhance ovarian function beyond the effects of BS alone through mechanisms such as augmented weight loss, improved insulin sensitivity, or reduced inflammation. Understanding the effects of post-BS exercise on ovarian function could hep clinicians make informed decisions to optimize fertility treatments.

Therefore, the aims of the EMOVAR trial were i) to investigate the effects of a 16-week supervised exercise intervention on the ovarian function (primary outcome: SHBG), weight loss, body composition, fitness, inflammation, and the cardiometabolic and metabolomic profiles in reproductive-aged women with severe obesity at week 16 and 1-year following BS; and (ii) to examine whether weight loss and changes in the secondary/exploratory outcomes are associated with changes in the ovarian function.

## METHODS

### Study design and participants

The EMOVAR study was a two-arm parallel-group, data analyst and outcome-assessor blinded, randomized trial. The study protocol is published ^[24]^. The patients who volunteered to participate were recruited, one by one, as they received their appointments for BS (approximately one moth before BS; supplementary figure S1) from the Bariatric Surgery Units of two hospitals in the same region between October 2019 to September 2021.

Reproductive-aged women (18-45 years) with a BMI of ≥ 40 kg/m^2^ (or BMI ≥ 35 with comorbidities), and a history of obesity for at least 5 years were included. The complete eligibility criteria are presented at supplementary table S1. At recruitment, the patients signed written informed consent both for the surgical procedure and study participation. The trial was approved by the Local Ethics Committee and adhered to the Consolidated Standards of Reporting Trials (CONSORT; supplementary table S2) ^[25]^.

### Sample size

The sample size was calculated for the primary outcome (i.e., SHBG) based on previous studies examining the effects of exercise in obese women ^[26]^. With an anticipated between-group difference of 10 nmol/L, a statistical power of 85%, a 5% alpha-error, and a potential dropout of 10%, we aimed to recruit a minimum of 40 women (at least 20 per group).

### Randomization, treatment allocation and blinding

A computer-generated simple randomization sequence (1:1 ratio) was performed to allocate participants either to BS + usual care (BS+UC) or BS + exercise (BS+EX). Individual allocations were held in sealed, opaque, and consecutively numbered envelopes. Each participant was randomized by a blinded nurse at medical discharge following BS, provided they met inclusion criteria, signed informed consent, and had performed baseline assessments before BS.

### Study treatments

The intervention period and complete data collection run from October 2019 through September 2022. The outcomes were assessed at baseline (3-12 days before surgery), at week 16 (end of the intervention), and 1 year. A comprehensive timeline of the design is presented in supplementary figure S1. The surgical techniques performed were either traditional or one anastomosis gastric bypass or laparoscopic sleeve gastrectomy (i.e., only when BMI≥50).

Both groups received standard healthy lifestyle recommendations and identical dietary guidance focused on healthy eating, in line with international post-BS guidelines ^[27]^. In addition to the usual care following BS, the BS+EX group undertook a 16-week exercise program that included three 1-hour individual training sessions per week, totaling 48 sessions. These sessions combined resistance and aerobic training, organized into four blocks to gradually increase complexity and intensity for each participant. The total exercise training volume was 180 min/week. The program took place at the sport facilities from the university and was supervised by exercise professionals with undergraduate degrees in Sports Sciences and at least 2 years of experience as personal trainers for individuals with obesity. The training program used in the EMOVAR trial has been designed by our research group following the Consensus on Exercise Reporting Template (CERT) and a comprehensive description of the intervention is published elsewere^[28]^. The sessions were structured as follows: 1) warm-up; 2) compensatory training; 3) resistance training; 4) aerobic training; and 5) cool down (supplementary table S3). The warm-up comprised 5 min of light aerobic exercise (50-65% of Heart Rate Reserve [HRR]) on a treadmill, while the cool down included 5 min of flexibility exercises. Compensatory training included core stability exercises to minimize the risk of injury during the subsequent resistance training exercises. For resistance training, exercises progressed in intensity based on the participant’s response to the exercise during the four phases: familiarization, phase 1, 2, and 3. During the first 4 weeks of familiarization, participants learned main movement patterns and performed weight-bearing and resistance training with elastic bands to ensure appropriate technique. From phase 1 onwards, exercises included whole-body movement patterns (squat, seated lat pull-down, bench press, seated low row, push press with dumbbells, and deadlift), progressing from 1 to 3 sets, from 12 to 6 repetitions per set, interspersed by 30-60 seconds of rest, and ranging from 50% to 75% of one-repetition maximum (1RM). Resistance training intensity was quantified by the level of effort scale (i.e., number of repetitions actually performed out of the maximum number of repetitions that could be performed with that load). Participants were instructed to perform the concentric phase with the maximum intended velocity ^[29]^. Aerobic training was performed on a treadmill after resistance training. The training volume progressed from 15 min (familiarization phase) to 25 min (phase 4), while the intensity ranged from 65 to 85% of HRR. Aerobic training intensity was determined using the Karvonen’s formula ^[30]^. Trainers monitored the training intensity using heart rate monitors (Polar V800) and the rate of perceived exertion (RPE) 0-10 scale (e.g., a value of 7 to 9 corresponds to 75-85% of HRR). If the target aerobic intensity (% of HRR) was not achieved by increasing speed (without running), the treadmill’s incline was adjusted accordingly.

To enhance adherence, weekly motivational messages and videos were sent via WhatsApp to each participant. Trainers recorded attendance, mood, adverse events, or compliance daily, and monitored RPE from exercise sessions to anticipate potential fatigue and make necessary adjustments.

### Outcome measures

The outcomes were measured in serum. At each visit, blood samples were collected at the hospital at 9:00 am on the first assessment day, following a minimum 12-hour fasting period. A volume of 20mL of blood was drawn, processed, frozen, and stored. The adverse events were recorded throughout the trial.

The primary outcome was SHBG analyzed by immunoassay using the Beckman Coulter kit (ref. A48617), with a maximum value of 200nmol/L and inaccuracy <7%. The secondary outcomes related to ovarian function comprised i) changes in anti-müllerian hormone (AMH), FSH, thyroxine (T4), thyrotropin (TSH), luteinizing hormone (LH), estradiol, prolactine (PRL), total testosterone and FAI (supplement 2, pp 4); ii) changes in oocyte count, ovarian volume, endometrial thickness and uterine artery mean pulsatility index (UtA-PI) obtained from a transvaginal ultrasound using Toshiba Xario ultrasound equipment (Toshiba Medical Systems Corporation, Japan; supplement 1 pp 4). Other secondary outcomes were changes in traditional markers of chronic inflammation (CRP, leptin, TNF-α) and cardiometabolic profile (glucose, insulin, HOMA-IR, and pulse wave velocity; supplement 3 pp 4-5). Weight loss and changes in body composition were assessed with bioelectrical impedance InBody 270 (InBody CoLtd, Seoul, South Korea). Total weight loss (in %) was calculated as (weight loss = [pre-operative weight – weight follow-up] / pre-operative weight) ^[31]^. Changes in physical fitness were assessed using the Bruce treadmill test (cardiorespiratory fitness) ^[32]^, 30-second chair stand test (lower body strength) ^[33]^, handgrip test (upper body strength) ^[34]^, and back-scratch test (flexibility) ^[33]^. Detailed descriptions of these tests are available in the supplementary material (supplement 4, pp. 5–6). Lastly, changes in self-reporting of food intake were assessed through the food frequency queestionnarie used in the PREDIMED study ^[35]^.

### Exploratory outcomes not pre-specified (Metabolomics)

A Nuclear Magnetic Resonance (NMR)-based metabolomic study was conducted on the serum samples with a Bruker Avance III 600 spectrometer equipped with a quadruple HCNP cryoprobe and a thermostated SampleJet autosampler at 300 ± 0.1K. . Several classes of metabolites were identified including carbohydrates (e.g., glucose), amino acids (AAs), tricarboxylic acid cycle metabolites (e.g., citric acid, pyruvic acid), fatty acids and ketone bodies (e.g., 3-hydroxybutyrate and acetate). Multivariate data analyses were applied to assess the development of predictive models and potential biomarkers. Detailed descriptions of the sample preparation, metabolite assignments, and the statistical analyses conduced for these outcomes are available in the supplementary material (supplement 5 pp 6-7), along with a comprehensive explanation of the statistical analyses conducted for these outcomes.

### Deviations from the original protocol

The DEXA scan (DMS Imaging, STRATOS dR) indicated in the original protocol was not available due to device-related technical issues and we had to use bioelectrical impedance.

### Statistical Analysis

Baseline descriptive statistics are presented as frequency and percentage for categorical outcomes and mean and SD for continuous variables. The comparability of the groups at baseline was checked. The between-group differences in the change from baseline to week 16 and from baseline to 1 year in the continuous outcomes were analysed with a linear mixed-effects model. All analyses were adjusted for the date of last menstruation period. The terms “group”, “time”, and “group × time interaction” were entered as fixed effects, and the individual as random effect. The primary analyses were per-protocol, as indicated in the original protocol, and we undertook sensitivity (i.e., intention-to-treat) analyses. We assessed whether changes in ovarian function could be predicted by weight loss and/or changes in secondary/exploratory outcomes within each group with forward stepwise regression. Since most outcomes did not follow a normal distribution, all values were log2-transformed for the following analyses. We calculated a fold change (FC) with the log2-transformed outcomes (i.e., log2 outcome at week 16 or 1-year post-BS / log2 outcome at baseline). Pearson correlation examined the association of changes in serum metabolites with changes in ovarian function-related outcomes. To account for the false discovery rate (FDR), all *p*-values were corrected by the two-stage step-up method of Benjamini-Hochberg ^[36]^. The statistical analyses were performed with SPSS v.25.0 (IMB Corporation, Chicago, IL, USA) and R-software, v.4.0.3 (R Foundation for Statistical Computing). The statistical significance was set at *p*<0.05.

## RESULTS

In the EMOVAR study, 46 participants were randomized (25 to BS+UC, 21 to BS+EX). A total of 44 patients (96%) underwent gastric bypass procedures and 2 (4%) underwent laparoscopic sleeve gastrectomy. One participant in the BS+UC group was lost to follow-up during the COVID pandemic. Three participants in the BS+EX did not adhere to the exercise program. In total, 42 participants (91%; 24 in BS+UC; 18 in BS+EX) were included in the per-protocol analyses (Figure 1). Supplementary figure S2 shows participants’ attendance and average exercise intensity per session (i.e., HRR and RPE). The average attendace was 88.1 ± 7.5%. No adverse events were reported, except one participant with Guillain-Barré syndrome in the BS+EX group (not related to the intervention) who discontinued participation. The baseline characteristics of the study participants are presented in Table 1.

**Figure 1.**
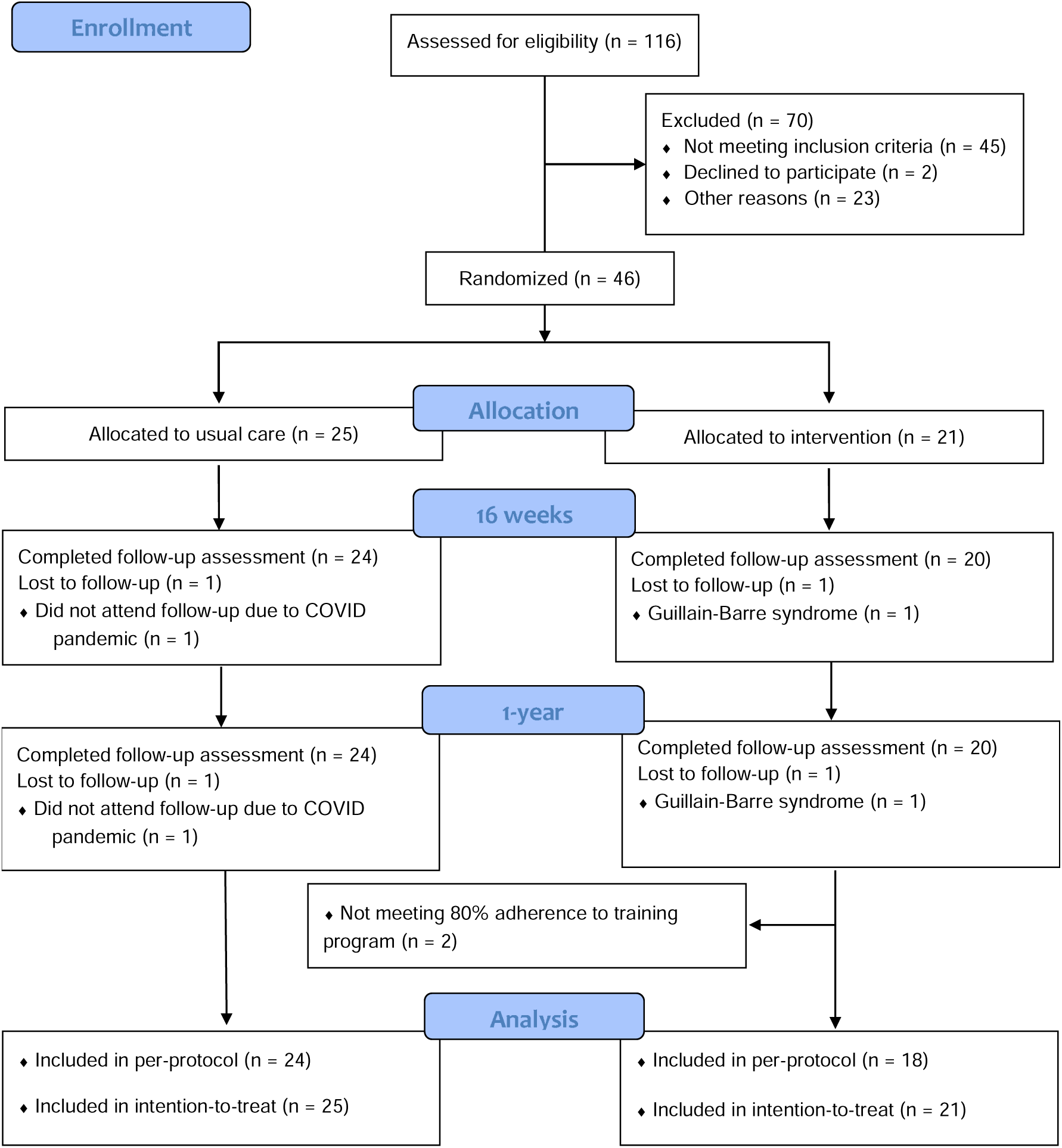
Participant enrollment in the EMOVAR study.

**Table 1.** Baseline Participant Characteristics.

| **Characteristics** | **All (n= 46)** | **Control group**<br>**(n=25)** | **Intervention group**<br>**(n=21)** |
| --- | --- | --- | --- |
| Age, mean (SD), y | 36.6 (6.9) | 36.6 (7.5) | 36.6 (6.4) |
| Marital status |  |  |  |
| Single | 24 (52.2%) | 12 (48.0%) | 12 (57.1%) |
| Married | 19 (41.3%) | 11 (44.0%) | 8 (38.1%) |
| Divorced | 3 (6.5%) | 2 (8.0%) | 1 (4.8%) |
| Occupational status |  |  |  |
| Employed | 36 (78.3%) | 19 (76.0%) | 17 (81.0%) |
| Unemployed | 9 (19.6 %) | 5 (20.0%) | 4 (19.0%) |
| Retired | 1 (2.2%) | 1 (4.0%) | 0 (0%) |
| Medical conditions |  |  |  |
| Type 2 diabetes | 2 (4.3%) | 1 (4.0%) | 1 (4.8%) |
| Dyslipidemia | 4 (8.7%) | 3 (12.0%) | 1 (4.8%) |
| Obstructive sleep apnea syndrome | 8 (17.4%) | 5 (20.0%) | 3 (14.3%) |
| Polycystic Ovary Syndrome | 1 (2.2%) | 1 (4.0%) | 0 (0%) |
| Menstrual status |  |  |  |
| Eumenorrhea | 40 (87.0%) | 22 (88.0%) | 18 (85.7%) |
| Amenorrhea | 6 (13.0%) | 3 (12.0%) | 3 (14.3%) |
| Medication |  |  |  |
| Antihypertensive | 1 (2.2%) | 0 (0%) | 1 (4.8%) |
| Statin | 10 (21.7%) | 6 (24.0%) | 4 (19.0%) |
| Oral antidiabetic | 0 (0%) | 0 (0%) | 0 (0%) |
| Insulin | 0 (0%) | 0 (0%) | 0 (0%) |
| Body height, mean (SD), cm | 162.1 (6.1) | 162.8 (6.2) | 161.3 (5.9) |
| Body weight, mean (SD), kg | 122.3 (17.1) | 125.3 (17.5) | 118.6 (16.4) |
| BMI | 46.6 (6.5) | 47.4 (7.3) | 45.5 (5.5) |
| Obesity class I (BMI 30 to 34.9) | 0 (0%) | 0 (0%) | 0 (0%) |
| Obesity class II (BMI 35 to 39.9) | 6 (13%) | 3 (12%) | 3 (14.3%) |
| Obesity class III (BMI ≥ 40) | 40 (87%) | 22 (88%) | 18 (85.7%) |
| Alcohol consumption |  |  |  |
| Never | 14 (30.4%) | 9 (36.0%) | 5 (23.8%) |
| Occasionally | 24 (52.2%) | 13 (52.0%) | 11 (52.4%) |
| Frequently | 7 (15.2%) | 2 (8.0%) | 5 (23.8%) |
| Daily | 1 (2.2%) | 1 (4.0%) | 0 (0%) |
| Tobacco consumption |  |  |  |
| Nonsmoker | 38 (82.6%) | 19 (76.0%) | 19 (90.5%) |
| Ex-smoker | 5 (10.9%) | 3 (12.0%) | 2 (9.5%) |
| Smoker | 3 (6.5%) | 3 (12.0%) | 0 (0%) |
| Anti contraceptive methods |  |  |  |
| None | 26 (56.5%) | 14 (56.0%) | 13 (61.9%) |
| Yes | 20 (43.5%) | 11 (44.0%) | 8 (38.1%) |
| Hirsutism (score) |  |  |  |
| Normal ($\leq 8$) | 0 (0%) | 0 (0%) | 0 (0%) |
| Mild (8 to 11) | 29 (63.0%) | 13 (52.0%) | 16 (76.2%) |
| Moderate (12 to 19) | 17 (37.0%) | 12 (48.0%) | 5 (23.8%) |
| Severe ($\geq 20$) | 0 (0%) | 0 (0%) | 0 (0%) |
| Surgery done |  |  |  |
| One anastomosis gastric bypass, n (%) | 42 (91.3%) | 22 (88.0%) | 20 (95.2%) |
| Gastric bypass, n (%) | 2 (4.3%) | 1 (4.0%) | 1 (4.8%) |
| Laparoscopic sleeve gastrectomy, n (%) | 2 (4.3%) | 2 (8.0%) | 0 (0%) |
| Hospital |  |  |  |
| Public | 11 (23.9%) | 8 (32.0%) | 3 (14.3%) |
| Private | 35 (76.1%) | 17 (68%) | 18 (85.7%) |

N (%), number of participants (percentage of participants within the group). SD, standard deviation BMI, body mass index.

### Exercise following BS improves relevant markers of ovarian function despite comparable benefits for traditional cardiometabolic risk markers in comparison to usual care

At week 16, the BS+EX increased the endometrial thickness (+3.7 mm, 95%CI 1.0 to 6.3; *p* = 0.008) and decreased the UtA-PI ((−1.3; 95%CI −2.1 to −0.5; *p=*0.003) compared to 0.8, 95% CI −1.6 to −0.1; *p* = 0.038) compared to BS+UC, although there were no effects on SHBG (+28.5 nmol/L; 95% CI −3.9 to 60.9; *p* = 0.084) or the other secondary outcomes related to ovarian function (Table 2). At 1-year, the BS+EX group significantly increased the serum levels of SHBG by 115% (+36.3 nmol/L; 95% CI 2.3 to 70.2; *p =* 0.037) and the oocyte count by 25.9% (+3.1 follicles; 95% CI 0.9 to 5.3; *p =* 0.007), and reduced the UtA-PI levels by 31.9% ((−1.3; 95%CI −2.1 to −0.5; *p=*0.003) compared to 1.3; 95% CI −2.1 to −0.5; *p =* 0.003) compared to BS+UC (Table 2).

**Table 2.** Changes in sex hormone levels and ovarian function after 16-week of supervised exercise intervention and after 1-year post bariatric surgery.

| End point | Bariatric surgery +<br>usual care (n=24) | Bariatric surgery +<br>exercise (n=18) | Mean difference<br>between groups |
| --- | --- | --- | --- |
| SHBG (nmol/L) |  |  |  |
| Baseline | 60.9 (38.1 to 83.7) | 43.3 (16.7 to 69.9) | - |
| Change at week 16 | 35.1 (10.1 to 60.1) | 63.6 (33.6 to 93.6) | 28.5 (-3.9 to 60.9) |
| Change at 1-year | 45.4 (19.6 to 71.3) | 81.7 (50.0 to 113.4) | **36.3 (2.3 to 70.2)\*** |
| AMH (ng/mL) |  |  |  |
| Baseline | 2.9 (1.7 to 4.1) | 3.1 (1.7 to 4.5) | - |
| Change at week 16 | -0.7 (-1.4 to 0.1) | -0.5 (-1.3 to 0.3) | 0.2 (-0.6 to 1.1) |
| Change at 1-year | -0.8 (-1.5 to 0.1) | -0.4 (-1.2 to 0.5) | 0.5 (-0.4 to 1.4) |
| FSH (mIU/mL) |  |  |  |
| Baseline | 7.4 (5.7 to 9.2) | 6.8 (4.8 to 8.8) | - |
| Change at week 16 | 0.1 (-2.7 to 2.9) | 0.5 (-2.8 to 3.9) | 0.4 (-3.1 to 4.0) |
| Change at 1-year | 0.2 (-2.7 to 3.1) | 0.3 (-3.2 to 3.8) | 0.1 (-3.6 to 3.8) |
| LH (mIU/mL) |  |  |  |
| Baseline | 10.1 (5.8 to 14.4) | 6.9 (1.9 to 11.9) | - |
| Change at week 16 | -3.3 (-10.5 to 3.9) | 0.6 (-8.0 to 9.1) | 3.8 (-5.3 to 13.0) |
| Change at 1-year | 2.1 (-5.3 to 9.4) | -0.2 (-9.1 to 8.7) | -2.3 (-11.7 to 7.2) |
| TSH (μIU/mL) |  |  |  |
| Baseline | 2.2 (1.6 to 2.8) | 1.8 (1.1 to 2.6) | - |
| Change at week 16 | -0.4 (-1.0 to 0.2) | -0.5 (-1.3 to 0.3) | -0.1 (-0.9 to 0.7) |
| Change at 1-year | -0.5 (-1.2 to 0.1) | -0.3 (-1.1 to 0.5) | 0.2 (-0.6 to 1.1) |
| Free T4 (ng/dL) |  |  |  |
| Baseline | 0.89 (0.84 to 0.94) | 0.94 (0.90 to 1.00) | - |
| Change at week 16 | -0.02 (-0.09 to 0.05) | -0.07 (-0.14 to 0.01) | -0.05 (-0.14 to 0.04) |
| Change at 1-year | -0.05 (-0.12 to 0.02) | -0.04 (-0.12 to 0.04) | -0.01 (-0.08 to 0.10) |
| Prolactin (ng/mL) |  |  |  |
| Baseline | 13.0 (10.4 to 15.5) | 12.2 (9.3 to 15.2) | - |
| Change at week 16 | -1.2 (-3.9 to 1.6) | -1.2 (-4.5 to 2.2) | 0.0 (-3.6 to 3.6) |
| Change at 1-year | -1.3 (-4.1 to 1.6) | -0.4 (-3.9 to 3.1) | 0.9 (-2.9 to 4.6) |
| Estradiol (pmol/L) |  |  |  |
| Baseline | 345.0 (141.0 to 549.0) | 344.0 (107.0 to 581.0) | - |
| Change at week 16 | 100.2 (-193.0 to 393.0) | 60.8 (-289.0 to 410.0) | -39.3 (-417.4 to 338.7) |
| Change at 1-year | 126.6 (-175.0 to 428.0) | 174.2 (-192.0 to 541.0) | 47.6 (-341.1 to 436.2) |
| Progesterone (ng/mL) |  |  |  |
| Baseline | 1.1 (-0.5 to 2.8) | 2.1 (0.2 to 4.0) | - |
| Change at week 16 | 1.0 (-1.5 to 3.5) | 0.7 (-2.3 to 3.7) | -0.3 (-3.5 to 3.0) |
| Change at 1-year | 2.0 (-0.6 to 4.6) | -0.5 (-3.6 to 2.6) | -2.5 (-5.8 to 0.9) |
| Testosterone (nmol/L) |  |  |  |
| Baseline | 1.5 (1.2 to 1.8) | 1.6 (1.3 to 1.9) | - |
| Change at week 16 | -0.3 (-0.6 to 0.02) | -0.4 (-0.7 to -0.01) | -0.1 (-0.5 to 0.3) |
| Change at 1-year | -0.3 (-0.7 to 0.04) | -0.4 (-0.8 to 0.01) | -0.0 (-0.4 to 0.4) |
| Androstendione (ng/mL) |  |  |  |
| Baseline | 1.4 (0.9 to 1.9) | 1.9 (1.3 to 2.4) | - |
| Change at week 16 | -0.2 (-0.8 to 0.4) | -0.1 (-0.8 to 0.6) | 0.1 (-0.7 to 0.9) |
| Change at 1-year | -0.1 (-0.7 to 0.5) | -0.04 (-0.8 to 0.7) | 0.1 (-0.8 to 0.9) |
| FAI |  |  |  |
| Baseline | 3.7 (2.9 to 4.6) | 4.1 (3.1 to 5.1) | - |
| Change at week 16 | -2.2 (-3.2 to -1.3) | -2.2 (-3.4 to -1.1) | 0.0 (-1.2 to 1.3) |
| Change at 1-year | -2.6 (-3.6 to -1.6) | -2.3 (-3.5 to -1.0) | 0.3 (-1.0 to 1.6) |
| DHEA-S (µg/dL) |  |  |  |
| Baseline | 157.0 (12.5 to 189.0) | 169.0 (132.0 to 206.0) | - |
| Change at week 16 | -14.5 (-37.7 to 8.7) | -23.5 (-51.5 to 4.5) | -9.0 (-39.1 to 21.1) |
| Change at 1-year | -31.0 (-55.0 to -7.0) | -28.9 (-58.5 to 0.8) | 2.1 (-29.6 to 33.8) |
| Oocyte count (No) |  |  |  |
| Baseline | 13.2 (10.0 to 16.3) | 11.9 (8.2 to 15.6) | - |
| Change at week 16 | -0.1 (-1.9 to 1.5) | 1.3 (-0.9 to 2.9) | 1.4 (-0.7 to 3.5) |
| Change at 1-year | -1.8 (-3.0 to 0.5) | 1.2 (1.1 to 2.9) | **3.1 (0.9 to 5.3)\*** |
| Endometrial thickness (mm) |  |  |  |
| Baseline | 7.8 (6.5 to 9.1) | 4.6 (3.0 to 6.1) | - |
| Change at week 16 | -1.7 (-3.8 to 0.4) | 2.0 (-0.5 to 4.5) | **3.7 (1.0 to 6.4)\*** |
| Change at 1-year | -0.7 (-2.8 to 1.4) | 1.7 (-0.9 to 4.3) | 2.4 (-0.4 to 5.1) |
| Right ovarian volumen (cm<sup>3</sup>) |  |  |  |
| Baseline | 5.3 (4.0 to 6.7) | 6.1 (4.5 to 7.7) | - |
| Change at week 16 | 1.2 (-0.4 to 2.9) | 0.1 (-1.9 to 2.1) | -1.1 (-3.3 to 1.0) |
| Change at 1-year | 1.9 (0.2 to 3.6) | -0.1 (-2.2 to 2.1) | -2.0 (-4.2 to 0.3) |
| Left ovarian volumen (cm<sup>3</sup>) |  |  |  |
| Baseline | 5.3 (3.5 to 7.1) | 4.8 (2.7 to 6.9) | - |
| Change at week 16 | -0.3 (-2.2 to 1.5) | 1.3 (-0.9 to 3.5) | 1.6 (-0.8 to 4.0) |
| Change at 1-year | 0.1 (-1.5 to 2.0) | 1.1 (-1.3 to 3.4) | 1.0 (-1.5 to 3.5) |
| Right ovary mean diameter (mm) |  |  |  |
| Baseline | 21.8 (20.0 to 23.6) | 22.1 (20.0 to 24.2) | - |
| Change at week 16 | 2.0 (-0.3 to 4.3) | 0.5 (-2.2 to 3.2) | -1.5 (-4.4 to 1.4) |
| Change at 1-year | 2.0 (-0.3 to 4.4) | 1.0 (-1.9 to 3.8) | -1.0 (-4.1 to 1.9) |
| Left ovary mean diameter (mm) |  |  |  |
| Baseline | 21.1 (18.9 to 23.3) | 20.3 (17.8 to 22.8) | - |
| Change at week 16 | -0.2 (-2.7 to 2.4) | 2.1 (-1.0 to 5.2) | 2.2 (-1.1 to 5.5) |
| Change at 1-year | -0.2 (-2.8 to 2.5) | 1.7 (-1.5 to 5.0) | 1.9 (-1.6 to 5.3) |
| UtA-PI |  |  |  |
| Baseline | 3.0 (2.5 to 3.4) | 4.0 (3.5 to 4.6) | - |
| Change at week 16 | 0.4 (-0.2 to 1.1) | -0.4 (-1.1 to 0.3) | **-0.9 (-1.6 to -0.1)\*** |
| Change at 1-year | -0.0 (-0.7 to 0.6) | -1.3 (-2.1 to 0.6) | **-1.3 (-2.1 to -0.5)\*\*** |

SHBG, sex hormone-binding globulin; AMH, anti-müllerian hormone; FSH, follicle stimulating hormone; LH, luteinizing hormone; TSH, thyroid-stimulating hormone; FAI, free androgen index; DHEA-S, dehydroepiandrosterone sulfate; UtA-PI, uterine artery mean pulsatility index;
\*, $p < 0.05$ time × group interaction; \*\*, $p < 0.01$ time × group interaction.

There were no between-group differences in weight loss, cardiometabolic (Table 3), or NMR-metabolites (supplementary figure S3) from baseline to week 16 or 1 year, nor from week 16 to 1 year (supplementary tables S4, S5). We observed that more than half (61%) of the NMR-metabolites exhibited a significant main effect of time (*p* ≤ 0.05) (Figure 2), but non-significant clusters or group × time interactions were observed (supplementary figure S3). The results of the primary analyses were consistent when replicated with BMI, age, menstrual status or anti contraceptive methods as potential confounders (data not shown).

**Figure 2.**
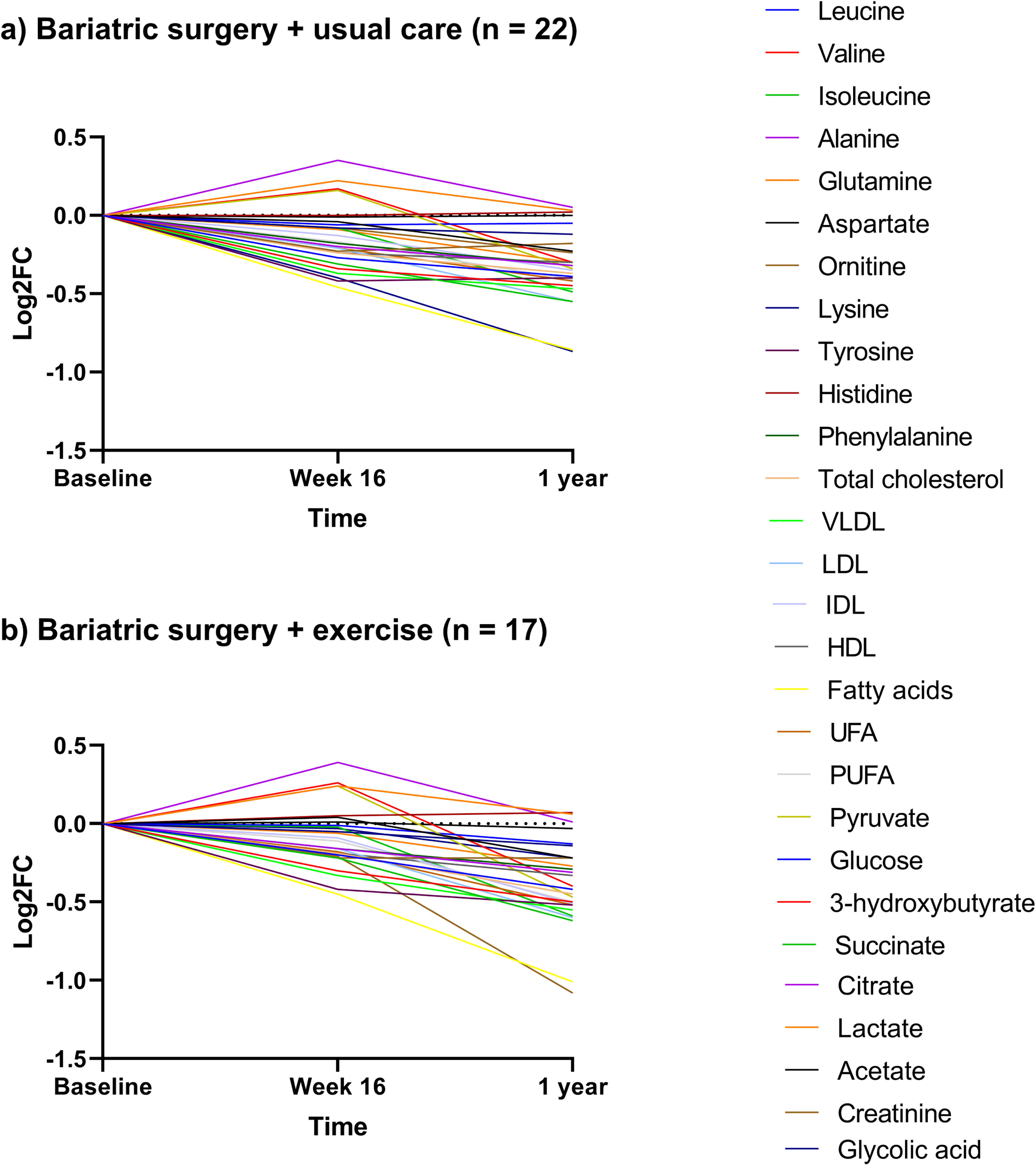
Dynamic changes in metabolomic profile throughout the longitudinal study. Metabolites are presented as Log2 fold change (Log2FC) relative to baseline.

**Table 3.** Changes in anthropometric, cardiometabolic and inflammatory profile after 16-week of supervised exercise intervention and after 1-year post bariatric surgery.

| End point | Bariatric surgery +<br>usual care (n=24) | Bariatric surgery +<br>exercise (n=18) | Mean difference<br>between groups |
| --- | --- | --- | --- |
| *Anthropometric profile* |  |  |  |
| Weight (kg) |  |  |  |
| Baseline | 126.1 (119.5 to 132.8) | 118.5 (110.7 to 132.8) | - |
| Change at week 16 | -28.4 (-32.1 to -24.8) | -29.7 (-34.4 to -25.1) | -1.3 (-6.2 to 3.6) |
| Change at 1-year | -47.3 (-51.2 to -43.5) | -45.8 (-50.7 to -40.8) | 1.6 (-3.6 to 6.8) |
| Weight loss (%) |  |  |  |
| Baseline | - | - | - |
| Change at week 16 | 22.0 (21.0 to 24.0) | 25.0 (23.0 to 28.0) | 2.0 (-0.4 to 5.0) |
| Change at 1-year | 38.0 (36.0 to 41.0) | 39.0 (36.0 to 42.0) | 1.0 (-2.2 to 4.2) |
| BMI (kg/m<sup>2</sup>) |  |  |  |
| Baseline | 47.7 (44.7 to 50.8) | 45.7 (43.0 to 48.4) | - |
| Change at week 16 | -10.7 (-12.1 to -9.3) | -11.3 (-13.0 to -9.5) | -0.6 (-2.4 to 1.2) |
| Change at 1-year | -18.0 (-19.4 to -16.5) | -17.4 (-19.3 to -15.5) | 0.6 (-1.2 to 2.6) |
| Body fat (%) |  |  |  |
| Baseline | 53.3 (50.9 to 55.6) | 51.8 (49.0 to 54.7) | - |
| Change at week 16 | -6.9 (-9.2 to -4.7) | -9.4 (-12.2 to -6.5) | -2.4 (-5.5 to 0.6) |
| Change at 1-year | -18.8 (-21.2 to -16.4) | -19.7 (-22.8 to -16.7) | -0.9 (-4.1 to 2.3) |
| FFM (kg) |  |  |  |
| Baseline | 58.7 (55.9 to 61.6) | 57.3 (53.9 to 60.7) | - |
| Change at week 16 | -6.9 (-8.6 to -5.3) | -6.1 (-8.2 to -3.9) | 0.9 (-1.3 to 3.1) |
| Change at 1-year | -8.2 (-10.0 to -6.4) | -7.7 (-10.0 to -5.5) | 0.5 (-1.9 to 2.8) |
| SMM (kg) |  |  |  |
| Baseline | 33.2 (31.5 to 34.9) | 32.2 (30.2 to 34.2) | - |
| Change at week 16 | -4.8 (-5.8 to 3.7) | -4.2 (-5.5 to 2.9) | 0.6 (-0.8 to 2.0) |
| Change at 1-year | -5.6 (-6.7 to 4.5) | -5.2 (-6.6 to 3.8) | 0.5 (-1.0 to 1.9) |
| *Cardiometabolic profile* |  |  |  |
| Glucose (mg/dL) |  |  |  |
| Baseline | 98.1 (93.6 to 102.6) | 98.0 (92.7 to 103.4) | - |
| Change at week 16 | -7.8 (-14.6 to -1.0) | -10.8 (-19.0 to -2.6) | -3.0 (-11.9 to 5.8) |
| Change at 1-year | -15.0 (-22.2 to -7.8) | -15.6 (-24.2 to -7.0) | -0.6 (-9.9 to 8.6) |
| Insulin (μU/mL) |  |  |  |
| Baseline | 18.7 (15.1 to 21.0) | 13.5 (10.0 to 17.0) | - |
| Change at week 16 | -9.5 (-13.2 to -5.7) | -6.9 (-11.4 to -2.3) | 2.6 (-2.3 to 7.5) |
| Change at 1-year | -13.0 (-17.0 to -9.0) | -8.3 (-13.1 to -3.5) | 4.7 (-0.5 to 9.9) |
| HOMA-IR |  |  |  |
| Baseline | 4.6 (3.7 to 5.4) | 3.5 (2.5 to 4.5) | - |
| Change at week 16 | -2.6 (-3.7 to -1.5) | -2.0 (-3.4 to -0.7) | 0.6 (-0.9 to 2.0) |
| Change at 1-year | -3.5 (-2.7 to -2.3) | -2.4 (-3.8 to -1.0) | 1.1 (-0.4 to 2.7) |
| Glycated hemoglobin<br>(mg/dL) |  |  |  |
| Baseline | 5.4 (5.2 to 5.5) | 5.4 (5.2 to 5.6) | - |
| Change at week 16 | -0.2 (-0.4 to -0.1) | -0.3 (-0.5 to -0.1) | -0.1 (-0.3 to 0.1) |
| Change at 1-year | -0.4 (-0.5 to -0.2) | -0.4 (-0.6 to -0.2) | -0.0 (-0.3 to 0.2) |
| Cortisol (μg/dL) |  |  |  |
| Baseline | 9.6 (8.0 to 11.1) | 10.0 (8.2 to 11.9) | - |
| Change at week 16 | -0.4 (-2.9 to 2.0) | 0.4 (-2.5 to 3.3) | 0.8 (-2.3 to 4.0) |
| Change at 1-year | 1.2 (-1.4 to 3.8) | 1.3 (-1.8 to 4.4) | 0.1 (-3.2 to 3.5) |
| HDL (mg/dL) |  |  |  |
| Baseline | 53.8 (50.1 to 57.4) | 50.9 (46.6 to 53.3) | - |
| Change at week 16 | -8.9 (-13.3 to 4.6) | -8.5 (-13.8 to 3.2) | 0.4 (-5.3 to 6.1) |
| Change at 1-year | 2.0 (-2.6 to 6.7) | 3.4 (-2.2 to 9.1) | 1.4 (-4.6 to 7.4) |
| **LDL (mg/dL)** |  |  |  |
| Baseline | 128.0 (113.5 to 142.0) | 115.8 (98.8 to 133.0) | - |
| Change at week 16 | -15.0 (-29.3 to 0.8) | -5.4 (-22.7 to 12.0) | 9.7 (-8.9 to 28.3) |
| Change at 1-year | -23.5 (-38.6 to 8.4) | -23.7 (-42.1 to 5.3) | -0.2 (-19.9 to 19.5) |
| **Triglycerides (mg/dL)** |  |  |  |
| Baseline | 121.7 (107.4 to 136.1) | 114.7 (97.6 to 131.8) | - |
| Change at week 16 | -21.5 (-42.4 to 0.5) | -30.0 (-55.4 to 4.6) | -8.6 (-35.5 to 18.4) |
| Change at 1-year | -54.3 (-76.5 to 32.1) | -48.9 (-75.5 to 22.3) | 5.4 (-22.9 to 33.7) |
| **Cholesterol (mg/dL)** |  |  |  |
| Baseline | 206.0 (191.0 to 222.0) | 190.0 (171.0 to 208.0) | - |
| Change at week 16 | -28.5 (-44.3 to 12.7) | -20.2 (-39.4 to 0.9) | 8.3 (-12.3 to 29.0) |
| Change at 1-year | -33.0 (-49.7 to 16.3) | -29.8 (-50.2 to 9.5) | 3.1 (-18.7 to 24.9) |
| **Aspartate transaminase (U/L)** |  |  |  |
| Baseline | 23.9 (19.8 to 28.0) | 23.7 (18.8 to 28.5) | - |
| Change at week 16 | 5.1 (-0.8 to 11.1) | 0.7 (-6.5 to 7.8) | -4.5 (-12.2 to 3.2) |
| Change at 1-year | 1.2 (-5.1 to 7.4) | -1.1 (-8.6 to 6.4) | -2.3 (-10.4 to 5.9) |
| **Alanine transaminase (U/L)** |  |  |  |
| Baseline | 27.4 (19.9 to 34.9) | 26.7 (17.8 to 35.6) | - |
| Change at week 16 | 6.2 (-4.0 to 16.5) | 1.5 (-10.9 to 13.9) | -4.8 (-17.9 to 8.4) |
| Change at 1-year | 0.9 (-9.8 to 11.8) | -0.3 (-13.3 to 12.8) | -1.2 (-15.1 to 12.7) |
| **Alkaline phosphatase (U/L)** |  |  |  |
| Baseline | 85.1 (74.5 to 95.7) | 84.3 (71.8 to 96.7) | - |
| Change at week 16 | 6.7 (-2.8 to 16.2) | 4.5 (-7.1 to 16.2) | -2.2 (-14.5 to 10.2) |
| Change at 1-year | 2.6 (-7.5 to 12.7) | 2.9 (-9.4 to 15.3) | 0.3 (-12.7 to 13.3) |
| **Creatinine (Mg/dL)** |  |  |  |
| Baseline | 0.7 (0.6 to 0.7) | 0.7 (0.6 to 0.7) | - |
| Change at week 16 | -0.1 (-0.1 to 0.01) | -0.1 (-0.1 to 0.03) | -0.0 (-0.1 to 0.04) |
| Change at 1-year | -0.1 (-0.1 to 0.01) | -0.1 (-0.1 to 0.01) | 0.0 (-0.1 to 0.1) |
| **Glomerular filtration rate (mL/min)** |  |  |  |
| Baseline | 113.0 (109.0 to 117.0) | 114.0 (109.0 to 118.0) | - |
| Change at week 16 | 5.5 (-1.3 to 9.7) | 6.0 (-0.9 to 11.1) | 0.5 (-5.0 to 6.0) |
| Change at 1-year | 5.8 (-1.3 to 10.2) | 4.5 (-0.9 to 9.9) | -1.3 (-7.1 to 4.5) |
| **Uric acid (mg/dL)** |  |  |  |
| Baseline | 5.8 (5.3 to 6.3) | 5.6 (5.0 to 6.2) | - |
| Change at week 16 | -0.6 (-1.2 to 0.01) | -0.9 (-1.7 to 0.2) | -0.3 (-1.1 to 0.4) |
| Change at 1-year | -1.5 (-2.2 to 0.9) | -1.5 (-2.3 to 0.8) | -0.0 (-0.8 to 0.8) |
| **Inflammation** |  |  |  |
| **TNF-α (pg/mL)** |  |  |  |
| Baseline | 7.67 (6.75 to 8.59) | 8.12 (7.05 to 9.19) | - |
| Change at week 16 | -0.74 (-2.28 to 0.80) | -2.65 (-4.48 to 0.83) | -1.91 (-3.89 to 0.06) |
| Change at 1-year | -4.95 (-6.57 to 3.34) | -4.87 (-6.77 to 2.97) | 0.08 (-1.98 to 2.14) |
| **Leptin (ng/mL)** |  |  |  |
| Baseline | 198.4 (181.7 to 215.1) | 176.3 (157.0 to 195.7) | - |
| Change at week 16 | -118.3 (-139.2 to 97.3) | -114.4 (-139.5 to 89.3) | 3.8 (-23.2 to 30.9) |
| Change at 1-year | -140.4 (-162.7 to 118.2) | -130.9 (-157.3 to 104.4) | 9.6 (-19.0 to 38.1) |
| **CRP (mg/dL)** |  |  |  |
| Baseline | 1.36 (1.07 to 1.65) | 0.88 (0.54 to 1.22) | - |
| Change at week 16 | -0.80 (-1.22 to 0.39) | -0.50 (-0.10 to 0.02) | 0.31 (-0.23 to 0.85) |
| Change at 1-year | -1.17 (-1.61 to 0.73) | -0.67 (-1.19 to 0.14) | 0.50 (-0.06 to 1.11) |
| Pulse wave velocity (m/s) |  |  |  |
| Baseline | 5.97 (5.69 to 6.26) | 6.11 (5.77 to 6.45) | - |
| Change at week 16 | -0.20 (-0.43 to 0.04) | -0.45 (-0.75 to 0.15) | -0.25 (-0.57 to 0.07) |
| Change at 1-year | -0.60 (-0.85 to 0.35) | -0.60 (-0.92 to 0.28) | 0.00 (-0.33 to 0.33) |
| **Physical fitness** |  |  |  |
| Handgrip strength (kg) |  |  |  |
| Baseline | 30.6 (28.9 to 32.3) | 29.5 (27.2 to 31.8) | - |
| Change at week 16 | -2.5 (-4.3 to -0.3) | -1.8 (-4.1 to 1.4) | 0.7 (-1.0 to 2.4) |
| Change at 1-year | -2.6 (-4.3 to -0.9) | -2.9 (-5.8 to 0.4) | -0.3 (-2.1 to 1.6) |
| Chair stand test (repetitions) |  |  |  |
| Baseline | 10.3 (9.36 to 11.2) | 12.4 (11.3 to 13.4) | - |
| Change at week 16 | 1.9 (0.7 to 3.3) | 2.5 (0.01 to 4.0) | 0.6 (-0.8 to 2.1) |
| Change at 1-year | 3.3 (1.8 to 5.0) | 2.8 (1.4 to 6.1) | -0.5 (0.8 to -2.0) |
| Bruce test (s) |  |  |  |
| Baseline | 327.0 (276.0 to 379.0) | 370.0 (301.0 to 438.0) | - |
| Change at week 16 | 134.6 (79.1 to 208.9) | 183.9 (47.5 to 246.4) | 49.3 (-36.0 to 134.6) |
| Change at 1-year | 229.7 (184.8 to 301.2) | 187.1 (29.4 to 300.6) | -42.5 (-132.1 to 47.0) |
| Back scratch (cm) |  |  |  |
| Baseline | -12.5 (-15.7 to -9.38) | -12.2 (-15.8 to -8.61) | - |
| Change at week 16 | 9.0 (6.5 to 14.3) | 8.3 (4.2 to 13.7) | -0.6 (-3.1 to 1.8) |
| Change at 1-year | 12.4 (8.4 to 15.7) | 10.9 (7.4 to 16.5) | -1.4 (-4.0 to 1.2) |

BMI, body mass index; FFM, fat free mass; SMM, skeletal muscle mass; HOMA-IR, homeostatic model assessment for insulin resistance; HDL, high-density lipoprotein; LDL, low-density lipoprotein; TNF-alpha, tumor necrosis factor alpha; CRP, C-reactive protein.

The intention-to-treat analyses corroborated results (supplementary tables S6, S7, S8, S9), except for a decrease in TNF-α ((−1.3; 95%CI −2.1 to −0.5; *p=*0.003) compared to 1.9 pg/mL, 95% CI −3.8 to 0.02) in the BS+EX at week 16 (supplementary table S7). There were no between-group differences in the frequency of food consumption either at week 16 or 1 year (supplementary tables S10 and S11).

### Changes in NMR-derived metabolites outperform traditional markers in predicting the changes in ovarian function

Overall, stepwise regression revealed that changes in the serum metabolites were stronger predictors of the changes in SHBG, oocyte count, and UtA-PI than the changes in traditional markers at week 16 (data not shown) and 1 year. This finding was observed both in the BS+EX (Table 4) and BS+UC groups (supplementary table S12). Specifically, the best predictors of the change in SHBG at 1-year post-BS differed across groups, with tyrosine and total cholesterol for BS+EX group (R^2^ 97%) and 3-hydroxybutyrate, leptin, and aspartate transaminase for the BS+UC group (R^2^ 90%).

### Changes in SHBG at 1 year are inversely related to changes in serum levels of amino acids only in the exercise group

The association of changes in serum metabolites and changes in markers of ovarian function at week 16 and 1-year are presented at supplementary figure S4 and Figure 3, respectively. The decrease in the panel of AAs (isoleucine, glutamine, ornithine, lysine, tyrosine, histidine, and phenylalanine) was associated with the increase in SHBG (*r_range_* from −0.56 to −0.67; all *p* < 0.05) only in the BS+EX group (Figure 3). None of the correlations in this section persisted after FDR correction.

**Figure 3.**
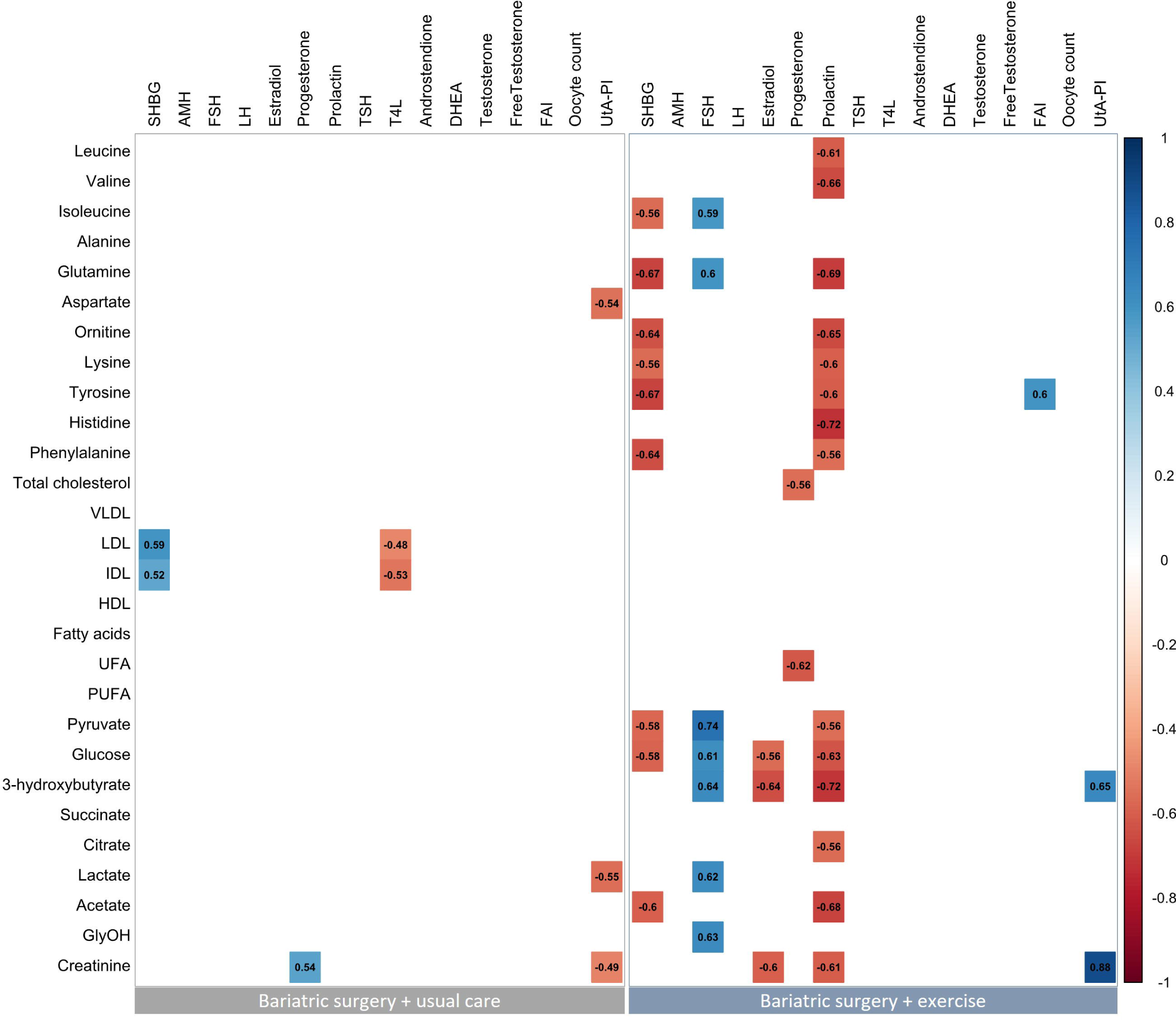
Pearson correlation analyses between changes in metabolites (derived from NMR) with changes in ovarian function at 1-year post bariatric surgery. Every box represents a significant correlation coefficient (all *p* < 0.05), whereas empty boxes represent no significant correlations. Significant after false discovery rate corrections.

## DISCUSSION

The main findings of the EMOVAR randomized trial suggest that a 16-week supervised exercise program improves relevant markers of ovarian function (specifically SHBG, oocyte count, and mean UtA-PI) at 1-year compared to usual care. Exercise did not enhance weight loss or provide additional benefits on body composition, physical fitness, chronic inflammation, and cardiometabolic and metabolomic profiles. Interestingly, we observed a decrease in serum AAs levels that was associated with an increase in SHBG at 1-year only in the BS+EX group. These results underline the potential of exercise to improve relevant markers of ovarian function in women undergoing BS, which might have clinical implications for the management of women with severe obesity wishing to conceive.

The EMOVAR exercise program increased the serum SHBG levels by 115% and the oocyte count by 25.9%, and reduced UtA-PI by 31.9% compared to usual care at 1-year. In similar cohorts, BS increased serum SHBG levels by ∼106-136% ^[37–39]^ and decreased serum testosterone levels by ∼25-42% ^[37,39]^ at 1-year. However, previous studies did not observe significant improvements in other sexual hormones like estradiol, progesterone, prolactin, LH or FSH, following either sleeve gastrectomy or gastric bypass procedures ^[37–40]^. These results aligns with the findings of the EMOVAR study, where 96% of participants underwent gastric bypass procedures. Of note, higher SHBG levels are linked to reduced hyperandrogenism in women ^[6]^, while higher oocyte count indicates increased ovarian reserve ^[41]^ and higher fertility rates ^[42]^. Additionally, higher UtA-PI is associated with lower risks of preeclampsia, gestational hypertensive disorders, and intrauterine growth restriction during pregnancy ^[43–45]^. Overall, our results suggest that exercise following BS might contribute to improve relevant markers of ovarian function. A recent network meta-analysis by our group ^[19]^ underscored that exercise is a crucial component of a holistic approach that also includes dietary and pharmacological strategies to improve the reproductive hormonal profile and restore ovulation in women with overweight or obesity, both with and without polycystic ovary syndrome. Based on the findings of the EMOVAR trial, we hypothesize that women undergoing BS wishing to conceive could benefit from combining exercise with pharmacological strategies (i.e., ovulation inducers) and a hypocaloric diet to further improve the reproductive hormonal profile and ovulation ^[19]^, which warrants further research.

The additional benefits of exercise on the ovarian function were not that evident at week 16. This lack of short-term effects of exercise may be attributed to the abrupt weight loss observed in both groups during this period (58% and 64% of the total weight loss at 1-year occurred within the first 16 weeks for BS+UC and BS+EX groups, respectively, Table 3). We know that BS affects the metabolomic ^[10]^ and gut microbiome ^[46]^ profiles, leading to changes in protein metabolism within the first 3 months after surgery ^[46]^. Similarly, BS reduces dyslipidemia rates by up to 77% within the first 6 months, with significant reductions in cholesterol by up to −31% and triglyceride levels up to −63% ^[47]^, in line with our results (Table 3). We observed that the improvement in insulin resistance, hyperandrogenism, and inflammation, which are all associated with ovarian dysfunction ^[2]^, were similar between groups at week 16. Hence, it is plausible to think that the metabolic advantages stemming from the removal, in average, of ∼22 kg of fat early after BS (i.e., ≥ 52% of the total fat loss occurred within the first 16 weeks; Table 3) may partially overshadow the immediate effects of almost any additional intervention, although this hypothesis requires to be investigated. Furthermore, Boppre et al. observed that exercise interventions of 12 weeks duration or more, starting beyond 6 months after BS, significantly reduce cardiometabolic risk factors post-surgery ^[21]^. Therefore, it is possible that longer exercise interventions may produce larger benefits following BS ^[21,48,49]^, especially if the exercise intervention starts shortly after surgery.

The metabolomic analyses provided further insights into the potential mechanisms behind the improvement of relevant markers of ovarian function through exercise at 1 year. We observed a significant decrease in AAs in both groups at week 16 and 1 year, a pattern previously observed following gastric bypass procedures ^[50]^. The reduction in the panel of several AAs was associated with increased SHBG only in the BS+EX group at 1 year. SHBG is a dimeric plasma glycoprotein composed by 402 AAs produced by the liver ^[51–53]^. Exercise can improve the uptake and utilization of AAs ^[54]^, while the liver plays a crucial role in regulating AAs levels in the bloodstream through processes such as synthesis, degradation, and conversion ^[55]^, Therefore, it is conceivable that the liver could allocate less resources towards AAs synthesis, allowing it to undertake other functions such as synthesizing more SHBG molecules, which would translate into a reduction of AAs in the blood under the exercise condition. This could partially explain the consistent and inverse correlations observed between the panel of AAs and SHBG in Figure 3 and suggest a potential mechanism through which exercise might increase SHBG levels in women following BS. However, this hypothesis remains speculative and future research is needed to address whether exercise interventions can alter SHBG isoforms and whether variations in AAs concentrations in cultured human hepatocytes or liver organoids influence SHBG synthesis and secretion.

### Limitations and strengths

The sex hormonal profile was not evaluated on the same day of the cycle for all women because it was not feasible from a logistic perspective and the primary outcome and several relevant secondary outcomes are not cycle-phase dependent. To address this limitation, all analyses were adjusted for the date of the last menstruation period, although residual confounding may have affected the analyses of cycle-dependent hormones such as FSH, LH, estradiol, or progesterone. We did not monitor the dietary intake throughout the intervention and potential group differences might have implied uncontrolled residual confounding.

However, in the EMOVAR trial, both groups were provided with the same dietary guidelines as part of the usual care, and no between-groups differences were observed in the frequency of food consumption at any time point, suggesting that exercise did not influence the diet, overall. It is plausible that a different type of exercise intervention might have yielded different results. For instance, Ruiz-Uribe et al. found that the effects of moderate intensity constant training (MIT) are related to improved oxidative capacity, while high-intensity interval training (HIIT) seems to enhance insulin sensitivity ^[56]^. However, the effects of HIIT on markers of ovarian function are currently unknown and high-intensity exercise might not be feasible at the early stages following BS, as was the case of the EMOVAR intervention. Future research should examine whether implementing HIIT at later stages after BS (e.g., after 3-4 months) could provide additional benefits on body composition, cardiometabolic risk factors, and markers of ovarian function. In addition, the EMOVAR trial was not designed to evaluate the effects of exercise on menstruation or menstrual irregularities but rather focused on the homonal profile and cardiometabolic risk factors. Therefore, future research should examine whether, in women with severe obesity and menstrual irregularities, longer exercise interventions with varying configurations following BS might improve the ovarian function, reduce menstrual irregularities, or restore menstruation. The EMOVAR trial has also strengths that need to be underlined. This is the first randomized trial evaluating the impact of exercise on the ovarian function after BS. Importantly, we undertook a comprehensive exercise intervention that is available open access ^[24]^, ensuring transparency and replicability in research settings and clinical practice, which is a significant strength considering previous research in the field ^[57]^. Finally, it is remarkable that the adherence to the exercise intervention (88.1%) can be considered highly satisfactory compared with previous studies ^[57,58]^, which enhances the confidence in the study results.

## CONCLUSION

The EMOVAR randomized trial suggests that a 16-week supervised exercise program improves relevant markers of ovarian function, such as SHBG, oocyte count, and UtA-PI, at 1 year compared to usual care. These benefits occurred despite comparable weight loss and changes in fitness, chronic inflammation, and cardiometabolic and NMR-derived metabolomic profiles between groups. The observed association between decreased serum AAs levels and increased SHBG levels only in the BS+EX group suggests that exercise might enhance the liver’s availabitliy of AAs and its capacity to sytenthize more SHBG molecules, warranting further research. Overall, these findings suggest that exercise following BS could be integrated into a holistic approach to enhance ovulation and fertility in women with severe obesity undergoing BS.

## Data Availability

All data produced in the present study are available upon reasonable request to the authors

## REFERENCES

[1] Blüher M. Obesity: global epidemiology and pathogenesis. Nat Rev Endocrinol 2019;15:288–98.

[2] Broughton DE, Moley KH. Obesity and female infertility: potential mediators of obesity’s impact. Fertil Steril 2017;107:840–7.

[3] Chen X, Xiao Z, Cai Y, et al. Hypothalamic mechanisms of obesity-associated disturbance of hypothalamic-pituitary-ovarian axis. Trends Endocrinol Metab 2022;33:206–17.

[4] Kalliala I, Markozannes G, Gunter MJ, et al. Obesity and gynaecological and obstetric conditions: umbrella review of the literature. BMJ 2017;359:j4511.

[5] Rachoń D, Teede H. Ovarian function and obesity—Interrelationship, impact on women’s reproductive lifespan and treatment options. Mol Cell Endocrinol 2010;316:172–9.

[6] Tchernof A, Toth MJ, Poehlman ET. Sex hormone-binding globulin levels in middle-aged premenopausal women. Associations with visceral obesity and metabolic profile. Diabetes Care 1999;22:1875–81.

[7] Weiss G, Goldsmith LT, Taylor RN, et al. Inflammation in Reproductive Disorders. Reprod Sci 2009;16:216–29.

[8] Alalwan AA, Friedman J, Park H, et al. US national trends in bariatric surgery: A decade of study. Surgery 2021;170:13–7.

[9] Sjöström L, Peltonen M, Jacobson P, et al. Bariatric Surgery and Long-term Cardiovascular Events. JAMA 2012;307:56.

[10] Vaz M, Pereira SS, Monteiro MP. Metabolomic signatures after bariatric surgery – a systematic review. Rev Endocr Metab Disord 2022;23:503–19.

[11] Major P, Matłok M, Pędziwiatr M, et al. Quality of Life After Bariatric Surgery. Obes Surg 2015;25:1703–10.

[12] Ezzat RS, Abdallah W, Elsayed M, et al. Impact of bariatric surgery on androgen profile and ovarian volume in obese polycystic ovary syndrome patients with infertility. Saudi J Biol Sci 2021;28:5048–52.

[13] Eid GM, McCloskey C, Titchner R, et al. Changes in hormones and biomarkers in polycystic ovarian syndrome treated with gastric bypass. Surg Obes Relat Dis 2014;10:787–91.

[14] Christ JP, Falcone T. Bariatric Surgery Improves Hyperandrogenism, Menstrual Irregularities, and Metabolic Dysfunction Among Women with Polycystic Ovary Syndrome (PCOS). Obes Surg 2018;28:2171–7.

[15] Magro DO, Geloneze B, Delfini R, et al. Long-term Weight Regain after Gastric Bypass: A 5-year Prospective Study. Obes Surg 2008;18:648–51.

[16] Harrison CL, Lombard CB, Moran LJ, et al. Exercise therapy in polycystic ovary syndrome: a systematic review. Hum Reprod Update 2011;17:171–83.

[17] Hakimi O, Cameron LC. Effect of Exercise on Ovulation: A Systematic Review. Sport Med 2017;47:1555–67.

[18] Ennour-Idrissi K, Maunsell E, Diorio C. Effect of physical activity on sex hormones in women: a systematic review and meta-analysis of randomized controlled trials. Breast Cancer Res 2015;17:139.

[19] Ruiz-González D, Cavero-Redondo I, Hernández-Martínez A, et al. Comparative efficacy of exercise, diet and/or pharmacological interventions on BMI, ovulation, and hormonal profile in reproductive-aged women with overweight or obesity: a systematic review and network meta-analysis. Hum Reprod Update 2024;30:472–87.

[20] da Silva ALG, Sardeli A V., André LD, et al. Exercise Training Does Improve Cardiorespiratory Fitness in Post-Bariatric Surgery Patients. Obes Surg 2019;29:1416–9.

[21] Boppre G, Diniz-Sousa F, Veras L, et al. Does Exercise Improve the Cardiometabolic Risk Profile of Patients with Obesity After Bariatric Surgery? A Systematic Review and Meta-analysis of Randomized Controlled Trials. Obes Surg 2022;32:2056–68.

[22] Coen PM, Tanner CJ, Helbling NL, et al. Clinical trial demonstrates exercise following bariatric surgery improves insulin sensitivity. J Clin Invest 2015;125:248–57.

[23] Carretero-Ruiz A, del Carmen Olvera-Porcel M, Cavero-Redondo I, et al. Correction to: Effects of Exercise Training on Weight Loss in Patients Who Have Undergone Bariatric Surgery: a Systematic Review and Meta-Analysis of Controlled Trials. Obes Surg 2019;29:3778–3778.

[24] Soriano-Maldonado A, Martínez-Forte S, Ferrer-Márquez M, et al. Physical Exercise following bariatric surgery in women with Morbid obesity. Medicine (Baltimore) 2020;99:e19427.

[25] Moher D, Hopewell S, Schulz KF, et al. CONSORT 2010 Explanation and Elaboration: updated guidelines for reporting parallel group randomised trials. J Clin Epidemiol 2010;63:e1–37.

[26] Aubuchon M, Liu Y, Petroski GF, et al. The impact of supervised weight loss and intentional weight regain on sex hormone binding globulin and testosterone in premenopausal women. Syst Biol Reprod Med 2016;62:283–9.

[27] Mechanick JI, Youdim A, Jones DB, et al. Clinical practice guidelines for the perioperative nutritional, metabolic, and nonsurgical support of the bariatric surgery patient--2013 update: cosponsored by American Association of Clinical Endocrinologists, the Obesity Society, and American Society for Metabolic & Bariatric Surgery. Obesity 2013;21:S1–27.

[28] Villa-González E, Barranco-Ruiz Y, Rodríguez-Pérez MA, et al. Supervised exercise following bariatric surgery in morbid obese adults: CERT-based exercise study protocol of the EFIBAR randomised controlled trial. BMC Surg 2019;19:127.

[29] Baena-Raya A, Rodríguez-Rosell D, González-Badillo JJ, et al. Resistance training intensity in individuals following bariatric surgery: the need for rigorous prescription and monitoring. Int J Obes 2024;48:1359–60.

[30] Karvonen MJ, Kentala E, Mustala O. The effects of training on heart rate; a longitudinal study. Ann Med Exp Biol Fenn 1957;35:307–15.

[31] Corcelles R, Boules M, Froylich D, et al. Total Weight Loss as the Outcome Measure of Choice After Roux-en-Y Gastric Bypass. Obes Surg 2016;26:1794–8.

[32] Bruce RA, Blackmon JR, Jones JW, et al. Exercising Testing in Adult Normal Subjects and Cardiac Patients. Ann Noninvasive Electrocardiol 2004;9:291–303.

[33] Rikli RE, Jones CJ. Development and Validation of a Functional Fitness Test for Community-Residing Older Adults. J Aging Phys Act 1999;7:129–61.

[34] Ruiz-Ruiz J, Mesa JLM, Gutiérrez A, et al. Hand size influences optimal grip span in women but not in men. J Hand Surg Am 2002;27:897–901.

[35] Fernández-Ballart JD, Piñol JL, Zazpe I, et al. Relative validity of a semi-quantitative food-frequency questionnaire in an elderly Mediterranean population of Spain. Br J Nutr 2010;103:1808–16.

[36] Benjamini Y, Krieger AM, Yekutieli D. Adaptive linear step-up procedures that control the false discovery rate. Biometrika 2006;93:491–507.

[37] Ernst B, Wilms B, Thurnheer M, et al. Reduced circulating androgen levels after gastric bypass surgery in severely obese women. Obes Surg 2013;23:602–7.

[38] Kjær MM, Madsbad S, Hougaard DM, et al. The impact of gastric bypass surgery on sex hormones and menstrual cycles in premenopausal women. Gynecol Endocrinol 2017;33:160–3.

[39] Paul R, Andersson E, Wirén M, et al. Health-Related Quality of Life, Sexuality and Hormone Status after Laparoscopic Roux-En-Y Gastric Bypass in Women. Obes Surg 2020;30:493–500.

[40] Snoek KM, Steegers-Theunissen RPM, Hazebroek EJ, et al. The effects of bariatric surgery on periconception maternal health: a systematic review and meta-analysis. Hum Reprod Update 2021;27:1030–55.

[41] Scantamburlo VM, Linsingen R V., Centa LJR, et al. Association between decreased ovarian reserve and poor oocyte quality. Obstet Gynecol Sci 2021;64:532–9.

[42] Dian T, Adhi P, Ar RSD, et al. Comparison of oocyte count, fertilization, and pregnancy rates in adenomyosis patients undergoing In Vitro Fertilization with short and long protocol controlled ovarian stimulation – Restospective study. Ann Med Surg 2022;82:104620.

[43] Bahado-Singh RO, Syngelaki A, Akolekar R, et al. Validation of metabolomic models for prediction of early-onset preeclampsia. Am J Obstet Gynecol 2015;213:530.e1–530.e10.

[44] Mönckeberg M, Arias V, Fuenzalida R, et al. Diagnostic Performance of First Trimester Screening of Preeclampsia Based on Uterine Artery Pulsatility Index and Maternal Risk Factors in Routine Clinical Use. Diagnostics 2020;10:182.

[45] Moros G, Boutsikou T, Fotakis C, et al. Insights into intrauterine growth restriction based on maternal and umbilical cord blood metabolomics. Sci Rep 2021;11:1–10.

[46] Penney NC, Yeung DKT, Garcia-Perez I, et al. Multi-omic phenotyping reveals host-microbe responses to bariatric surgery, glycaemic control and obesity. Commun Med 2022;2:1–18.

[47] Buchwald H, Buchwald JN, McGlennon TW. Systematic review and meta-analysis of medium-term outcomes after banded Roux-en-Y gastric bypass. Obes Surg 2014;24:1536–51.

[48] Boppre G, Diniz-Sousa F, Veras L, et al. Can exercise promote additional benefits on body composition in patients with obesity after bariatric surgery? A systematic review and meta-analysis of randomized controlled trials. Obes Sci Pract 2022;8:112–23.

[49] Coen PM, Menshikova E V., Distefano G, et al. Exercise and Weight Loss Improve Muscle Mitochondrial Respiration, Lipid Partitioning, and Insulin Sensitivity After Gastric Bypass Surgery. Diabetes 2015;64:3737–50.

[50] West KA, Kanu C, Maric T, et al. Longitudinal metabolic and gut bacterial profiling of pregnant women with previous bariatric surgery. Gut 2020;69:1452–9.

[51] UniProt. UniProt. 2024. SHBG_HUMAN

[52] Avvakumov G V., Cherkasov A, Muller YA, et al. Structural analyses of sex hormone-binding globulin reveal novel ligands and function. Mol Cell Endocrinol 2010;316:13–23.

[53] Simó R, Sáez-López C, Barbosa-Desongles A, et al. Novel insights in SHBG regulation and clinical implications. Trends Endocrinol Metab 2015;26:376–83.

[54] Egan B, Zierath JR. Exercise metabolism and the molecular regulation of skeletal muscle adaptation. Cell Metab 2013;17:162–84.

[55] Biolo G, Maggi SP, Williams BD, et al. Increased rates of muscle protein turnover and amino acid transport after resistance exercise in humans. Am J Physiol - Endocrinol Metab 1995;268:514–20.

[56] Ruíz-Uribe M, Enríquez-Schmidt J, Monrroy-Uarac M, et al. Moderate-Intensity Constant and High-Intensity Interval Training Confer Differential Metabolic Benefits in Skeletal Muscle, White Adipose Tissue, and Liver of Candidates to Undergo Bariatric Surgery. J Clin Med 2024;13:3273.

[57] Baena=Raya A, Martínez=Rosales E, Ruiz=González D, et al. Exercise interventions following bariatric surgery are poorly reported: A systematic review and a call for action. Obes Rev 2024;25:e13758.

[58] Soriano-Maldonado A, Villa-González E, Ferrer-Márquez M, et al. Replicability of exercise programs following bariatric surgery. Atherosclerosis 2018;278:330–1.

