## Supplementary appendix for "EXERCISE IN WOMEN UNDERGOING BARIATRIC SURGERY: IMPACT ON THE OVARIAN FUNCTION AND CARDIOMETABOLIC RISK FACTORS – THE EMOVAR STUDY"

**Table of contents**

Supplementary material and methods.

1. Transvaginal ultrasound
2. Secondary outcomes related to ovarian function
3. Markers of chronic inflammation and cardiometabolic profile
4. Physical fitness
    3.1 Bruce test
    3.2 30-s chair stand test
    3.3 Handgrip test
    3.4 Back scratch
5. Metabolomic profile

5.1.Nuclear magnetic resonance (NMR) experiments

5.2. NMR metabolite assignments

5.3. Statistical analysis

Supplementary results

*Supplementary tables (in order of appearance in text)*

Table supplementary S1: Eligibility criteria for participants.

Table supplementary S2: CONSORT checklist

Table supplementary S3: Training periodization of the EMOVAR trial

*Per-protocol analyses*

Table supplementary S4: Changes from week 16 to 1-year in sex hormone levels using per-protocol approach.

Table supplementary S5: Changes from week 16 to 1-year in secondary outcomes using per-protocol approach.

*Intention-to-treat analyses*

Table supplementary S6: Changes in sex hormonal profile using intention-to-treat approach.

Table supplementary S8: Changes from week 16 to 1-year in sex hormone levels using intention-to-treat approach.

Table supplementary S7: Changes in secondary outcomes using intention-to-treat approach.

Table supplementary S9: Changes from week 16 to 1-year in secondary outcomes using intention-to-treat approach.

Table supplementary S10: Changes in the food frequency questionnaire using per-protocol approach.

Table supplementary S11: Changes in the food frequency questionnaire using intention-to-treat approach.

Table supplementary S12: Forward stepwise regression models for the usual care group at 1-year.

*Supplementary figures (in order of appearance in text)*

Figure supplementary S1: Graphical representation of the EMOVAR randomized trial.

Figure supplementary S2: Summary of participants assistance and intensity during the exercise intervention.

Figure supplementary S3: Principal component analysis for the metabolomic profile across intervention end points.

Figure supplementary S4: Heatmap analysis at week 16.

### Supplementary Material and Methods

#### 1. Transvaginal ultrasound.

The oocyte count, ovarian volume, endometrial thickness, and uterine arterial pulsatility index were obtained using Toshiba Xario ultrasound equipment (Toshiba Medical Systems Corporation, Japan) equipped with a 7 MHz curved endovaginal transducer.

For the oocyte count, we recorded the total number of follicles between 2 and 8 mm in size displayed on both ovaries. To assess ovarian volume, the longitudinal, oblique, and transverse axes were measured graphically, and the ovarian volume of each ovary was calculated using the formula: volume = D1 × D2 × D3 × 0.52 [1], where D represents the longitudinal, oblique and transverse axes, respectively.

The thickness of the endometrium was measured in millimeters and displayed ultrasonically in a longitudinal section of the uterus.

To assess the uterine pulsatility index, the 7MHZ endovaginal transducer was positioned paramedially to the uterine cervix at the level of the inner cervical opening.

For selective identification, the vessel was identified with color Doppler using high-speed scales (between 30 and 50 cm/s). The insertion angle for the measurement had to be less than 45º. Three or more waves of similar characteristics were obtained for the measurement. The size of the Doppler sample was equivalent to the artery's diameter and placed in the vessel's center. The pulsatility index was then calculated.

#### 2. Secondary outcomes related to ovarian function

The secondary outcomes related to ovarian function comprised serum markers of anti-müllerian hormone [AHM, ng/mL; kit's ref. B13127], FSH [mIU/mL; kit's ref. 33520], thyroxine [T4, ng/dL; kit's ref. 33880], thyrotropin [TSH, µIU/mL; kit's ref. B63284], luteinizing hormone [LH, mIU/mL; kit's ref. 33510], estradiol [E2, pmol/L; kit's ref. B84493], prolactine [PRL, ng/mL; kit's ref. 33530], total testosterone [T, nmol/L; kit's ref. 33560] and FAI [calculated as total testosterone/SHBG ratio]).

#### 3. Markers of chronic inflammation and cardiometabolic profile.

Tumor necrosis factor-alpha (TNFα; pg/mL), and leptin (pg/mL) were measured by immunoassay using specific kits according to manufacturer's protocols: alpha Human ELISA Kit (Thermo Fisher Scientific; KHC3011) and Leptin Human ELISA Kit (Thermo Fisher Scientific; KAC2281), respectively.

High-sensitivity C-reactive protein (CRP; mg/L) levels were assessed by immunoturbidimetric analysis using the Beckman Coulter kit, with a detection limit of 80 mg/L and a coefficient of variation < 1 mg/L.

According to manufacturer's protocols, glucose and insulin were measured by immunoassay using Cobas kit. The HOMA (homeostasis model assessment of insulin resistance) was calculated using the formula ([insulin, Miu/L] × [glucose, mg/Dl]) / 405 [2].

Pulse wave velocity was assessed using the Mobil-O-Graph 24-hour pulse wave analysis monitor (IEM GmbH, Stolberg, Germany) [3]. This device utilizes oscillometry recorded by a blood pressure sleeve positioned on the brachial artery. The pulse wave velocity assessment was conducted while seated, following a 5-minute rest period, in a room with controlled temperature [4].

### 4. Physical fitness

#### 4.1. Bruce test

Cardiorespiratory fitness was evaluated using the Bruce submaximal treadmill protocol [5], consisting of five stages, each lasting 3 minutes. The stages were as follows: stage 1 at 2.7 km/h with a 10% inclination, stage 2 at 4 km/h with a 12% inclination, stage 3 at 5.5 km/h with a 14% inclination, stage 4 at 6.8 km/h with a 16% inclination, and stage 5 at 8 km/h with an 18% inclination. The test ended when the participant reached 85% of their estimated maximum heart rate (HRmax), calculated using the formula by Tanaka et al. [6]. Maximal oxygen uptake ($VO_{2max}$) in METs is estimated based on time to exhaustion.

#### 4.2. 30-s chair stand test

The 30-s chair stand test was performed to assess lower body strength. The test started with the participant seated in a chair of approximately 17 centimeters in height, maintaining a straight back and feet flat on the floor. The participant crossed their arms at the wrists, holding them against the chest. Upon the signal "go," the participant rises to a full standing position and then returns to a fully seated position, as many times as possible within a 30-s time frame. A researcher monitors for proper technique throughout. The final score is the total number of stands executed correctly within the 30-s period [7].

#### 4.3. Handgrip strength

The handgrip test was performed to assess upper body strength with a TKK 5101 Grip-D digital dynamometer (precision of 0.1 kg). Subjects maintained a standard bipedal position throughout the test, extending their arms fully and avoiding contact between the dynamometer and any part of their body except the hand being measured. Each subject performed two trials with each hand, alternating between hands. We used the following mathematical equation to determine each participant’s optimal grip span: $y = x/5 + 1.5$ cm, where x is the hand size (maximal width between first and firth finger measured in centimeters, and y is the optimal grip span at which the dynamometer should be settled before the test) [8]. Two trials were conducted for each arm. The average of these trials was used for further analyses.

#### 4.4. Back-scratch test

The back-scratch test was performed to assess the shoulders’ range of motion. Standing upright, the participant placed one hand behind the same-side shoulder with the palm facing the back and fingers fully extended, reaching down the middle of the back (elbow pointed upward). Simultaneously, the other hand was placed behind the back with the palm facing outward, reaching upward to try to touch or overlap the extended fingers of both hands. Pulling fingers together is prohibited. The measurement was taken in centimeters, representing the overlap or distance between the tips of the middle fingers. A negative score indicates a gap between the fingers, while a positive score indicates an overlap [7]. Two trials were conducted for each arm. The average of these trials was used for further analyses.


[1] Pfeifer S et al. Fertil Steril. 2015;103:e9–17. https://doi.org/10.1016/j.fertnstert.2014.12.093.

[2] Turner RC et al. Metabolism. 1979;28:1086–96. https://doi.org/10.1016/0026-0495(79)90146-X.

[3] Wei W et al. Blood Press Monit. 2010;15:225–8. https://doi.org/10.1097/MBP.0b013e328338892f.

[4] Townsend RR et al. Hypertension. 2015;66:698–722. https://doi.org/10.1161/HYP.0000000000000033.

[5] Bruce RA et al. Ann Noninvasive Electrocardiol. 2004;9:291–303. https://doi.org/10.1111/j.1542-474X.2004.93003.x.

[6] Tanaka H et al. J Am Coll Cardiol. 2001;37:153–6. https://doi.org/10.1016/S0735-1097(00)01054-8.

[7] Rikli RE et al. J Aging Phys Act. 1999;7:129–61. https://doi.org/10.1123/japa.7.2.129.

[8] Ruiz-Ruiz J et al. J Hand Surg Am. 2002;27:897–901. https://doi.org/10.1053/jhsu.2002.34315.


### 5. Metabolomic profile

#### 5.1. Sample preparation

Prior to preparation, 114 serum samples were thawed at room temperature. Then, 300 µL of every supernatant were mixed with 300 µL of D2O KH2PO4 buffer 0.075 M (pH 7.4) containing the sodium salt of 3-(trimethylsilyl) propionic-2,2,3,3-d4 acid (TSP, 0.1%, w/v) and sodium azide (NaN3, 90 µM) as an enzyme inhibitor. 500 µL of each mixture was transferred to 5 mm NMR tubes.

#### 5.2. NMR acquisition and metabolite assignment

A Bruker Avance III 600 MHz spectrometer operating at a frequency of 600.13 MHz along with a thermostatted SampleJet autosampler of 500 positions and a 5 mm QCI quadruple resonance pulse field gradient cryoprobe was used in order to record the 1H NMR experiments. These spectra were obtained at $300 \pm 0.1$ K and acquired by using the Carr-Purcell-Meiboom-Gill (CPMG) pulse sequence with water presaturation pulse for its suppression (Bruker 1D cpmgpr1d) to attenuate broad signals in spectra from macromolecules and/or proteins. Samples were measured without rotation, using 16 dummy scans and 32 scans. Acquisition parameters were set as follows: size of FID (TD)

= 32K, spectral width (SW) = 22.0 ppm, acquisition time (AQ) = 1.24 s, relaxation delay (D1) = 3 s, receiver gain (RG) = 203 and FID resolution (FIDRES) = 0.81 Hz. Acquisition and processing of spectra were carried out with TOPSPIN software (3.6.3 version). The spectrometer transmitter was locked to D2O frequency, and the spectra were automatically phased, baseline-corrected and calibrated to the TSP signal at 0 ppm. Metabolite assignments of these serum samples were accomplished through information derived from 2D NMR experiments (homo- and heteronuclear experiments) corresponding to 1H−1H (TOCSY) and 1H−13C (HSQC and HMBC), along with the use of some NMR databases (Chenomx and HMDB) and the collected literature. The TOCSY spectrum was acquired using a D1 of 1.0 s, a spectral width of 7201.595 Hz in both dimensions (F1 and F2) and a receiver gain of 203. Also, this experiment was processed using the sine-bell function (SSB = 2.0). The HSQC spectrum was acquired using a D1 of 1.5 s, with a spectral width of 7211.538 Hz at F2 and 34710.770 Hz at F1. It was also processed using a sine-bell function (SSB = 2.0). The HMBC spectrum was acquired using a D1 of 1.0 s, with a spectral width of 7812.500 Hz in F2 and 37730.041 Hz in F1. Quantification of metabolites was performed relative to the internal standard (TSP) by integrating peak areas from isolated signals.

#### 5.3. Statistical analysis

Each NMR spectrum was divided into 0.04 ppm chemical shift bins from δH 0.2 to 10.0 ppm using AMIX 3.9.15 (Bruker BioSpin GmbH, Rheinstetten, Germany), and buckets were obtained by integrating the corresponding spectral areas. Region of δH 4.92−4.76 ppm, which contains residual signals of H2O suppression, was excluded from the bucketing and analysis. Normalization was performed on the data prior to statistical analyses, by scaling the intensity of individual peaks to the total intensity of the full spectra excepting the mentioned regions. Multivariate data analysis was performed on the data using SIMCA-P software (v. 17.0, Umetrics). Exploratory Principal Component Analysis (PCA) were applied and scaled to Unit variance. Scores plots were generated for these models. Pearson correlation analyses were performed for the obtained metabolites depending on the time period (baseline, week 16 and 1 year) to analyze whether there was any association with the exercise and usual care groups. These statistical analyses were applied through GraphPad Prism v10.1.0.

**Table S1.** Eligibility criteria for participants.

**Inclusion criteria**

- Women of childbearing age (between 18 and 45 years old).
- Body mass index ≥40 or 35 kg/m2 with comorbidities.
- Acceptable surgical risk (defined by the anaesthetist’s approval).
- Obesity maintained for at least 5 years.
- Failure of previous treatments.
- Signed informed consent for surgical treatment.
- Do not present contraindications for supervised physical exercise
- Residing in Almería capital∗ or (alternatively) presenting a willingness/predisposition to move and attend training sessions 3 times a week for 16 weeks (if assigned to the exercise group).

**Exclusion criteria**

- Serious psychiatric disorders such as schizophrenia, personality disorders, eating disorders, untreated depression or suicidal tendencies.
- Neurological disorders that may interfere with physical exercise.
- Adrenal or thyroid pathology that may be the cause of obesity.
- Uncontrolled addiction to alcohol or drugs.
- Presence of hysterectomy and/or prior adnexectomy.
- Active inflammatory or infectious diseases.

**Table S2.** CONSORT 2010 checklist of information to include when reporting a randomized trial.

**CONSORT 2010 checklist of information to include when reporting a randomised trial***

| Section/Topic | Item No | Checklist item | Reported on page No |
|---|---|---|---|
| **Title and abstract** | | | |
| | 1a | Identification as a randomised trial in the title | 1 |
| | 1b | Structured summary of trial design, methods, results, and conclusions (for specific guidance see CONSORT for abstracts) | 1 |
| **Introduction** | | | 2-3 |
| Background and objectives | 2a | Scientific background and explanation of rationale | |
| | 2b | Specific objectives or hypotheses | 3 |
| **Methods** | | | |
| Trial design | 3a | Description of trial design (such as parallel, factorial) including allocation ratio | 3-4 |
| | 3b | Important changes to methods after trial commencement (such as eligibility criteria), with reasons | NA |
| Participants | 4a | Eligibility criteria for participants | 4 |
| | 4b | Settings and locations where the data were collected | 3-4 |
| Interventions | 5 | The interventions for each group with sufficient details to allow replication, including how and when they were actually administered | 4-5 |
| Outcomes | 6a | Completely defined pre-specified primary and secondary outcome measures, including how and when they were assessed | 5-6 |
| | 6b | Any changes to trial outcomes after the trial commenced, with reasons | 6 |
| Sample size | 7a | How sample size was determined | 4 |

| | | | |
|---|---|---|---|
| | 7b | When applicable, explanation of any interim analyses and stopping guidelines | NA |
| Randomisation: | | | |
| Sequence generation | 8a | Method used to generate the random allocation sequence | 4 |
| | 8b | Type of randomisation; details of any restriction (such as blocking and block size) | 4 |
| Allocation concealment mechanism | 9 | Mechanism used to implement the random allocation sequence (such as sequentially numbered containers), describing any steps taken to conceal the sequence until interventions were assigned | 4 |
| Implementation | 10 | Who generated the random allocation sequence, who enrolled participants, and who assigned participants to interventions | 3 |
| Blinding | 11a | If done, who was blinded after assignment to interventions (for example, participants, care providers, those assessing outcomes) and how | 3 |
| | 11b | If relevant, description of the similarity of interventions | 4 |
| Statistical methods | 12a | Statistical methods used to compare groups for primary and secondary outcomes | 6 |
| | 12b | Methods for additional analyses, such as subgroup analyses and adjusted analyses | 8 |
| **Results** | | | |
| Participant flow (a diagram is strongly recommended) | 13a | For each group, the numbers of participants who were randomly assigned, received intended treatment, and were analysed for the primary outcome | 9 |
| | 13b | For each group, losses and exclusions after randomisation, together with reasons | 9 |
| Recruitment | 14a | Dates defining the periods of recruitment and follow-up | 9 |
| | 14b | Why the trial ended or was stopped | 9 |

| | | | |
|---|---|---|---|
| Baseline data | 15 | A table showing baseline demographic and clinical characteristics for each group | Table 1 |
| Numbers analysed | 16 | For each group, number of participants (denominator) included in each analysis and whether the analysis was by original assigned groups | 9 |
| Outcomes and estimation | 17a | For each primary and secondary outcome, results for each group, and the estimated effect size and its precision (such as 95% confidence interval) | 9-10; Tables 2-3 |
| | 17b | For binary outcomes, presentation of both absolute and relative effect sizes is recommended | NA |
| Ancillary analyses | 18 | Results of any other analyses performed, including subgroup analyses and adjusted analyses, distinguishing pre-specified from exploratory | 9-10 |
| Harms | 19 | All important harms or unintended effects in each group (for specific guidance see CONSORT for harms) | 9 |
| **Discussion** | | | |
| Limitations | 20 | Trial limitations, addressing sources of potential bias, imprecision, and, if relevant, multiplicity of analyses | 13 |
| Generalisability | 21 | Generalisability (external validity, applicability) of the trial findings | 11-13 |
| Interpretation | 22 | Interpretation consistent with results, balancing benefits and harms, and considering other relevant evidence | 11-13 |
| **Other information** | | | |
| Registration | 23 | Registration number and name of trial registry | 3 |
| Protocol | 24 | Where the full trial protocol can be accessed, if available | 3 |
| Funding | 25 | Sources of funding and other support (such as supply of drugs), role of funders | 3 |

**Table S3.** Training periodization of the EMOVAR trial.

| | **Phases** | **Familiarization phase** | | | | | | | | | | | | **Phase 1** | | | | | | | | | | | |
|---|---|---|---|---|---|---|---|---|---|---|---|---|---|---|---|---|---|---|---|---|---|---|---|---|---|
| | **Weeks** | **1** | | | **2** | | | **3** | | | **4** | | | **5** | | | **6** | | | **7** | | | **8** | | |
| | **Session** | ***1*** | ***2*** | ***3*** | ***4*** | ***5*** | ***6*** | ***7*** | ***8*** | ***9*** | ***10*** | ***11*** | ***12*** | ***13*** | ***14*** | ***15*** | ***16*** | ***17*** | ***18*** | ***19*** | ***20*** | ***21*** | ***22*** | ***23*** | ***24*** |
| | **Warm-up** | 5 min of low intensity aerobic activity (50-65% HRR) on Treadmill | | | | | | | | | | | | | | | | | | | | | | | |
| | **Compensatory training *** | - | | | | | | Core stability and stabiliser muscles (Level A) | | | | | | Core stability and stabiliser muscles (Level A-B) | | | | | | | | | | | |
| **Resistance training** | **Objective** | Initial adaptations to strength training | | | | | | | | | | | | Metabolic stress | | | | | | | | | | | |
| | **Type** | Learning exercises of movement patterns | | | | | | Learning exercises of movement patterns, weight-bearing and strength training with elastic bands | | | | | | 6 main resistance exercises focused in major muscle groups | | | | | | | | | | | |
| | **Sets** | 3 | | | | | | 2 | | | | | | 1 | 1 | 1 | 1 | 1 | 1 | 2 | 2 | 2 | 2 | 2 | 2 |
| | **Repetitions** | 8-10 | | | | | | 5-7 | | | | | | 12 | 12 | 12 | 12 | 12 | 12 | 12 | 12 | 12 | 12 | 12 | 12 |
| | **Intensity (LE)** | Weight-bearing | | | | | | Thera-Band RPE scale | | | | | | (24) | (24) | (24) | (24) | (24) | (24) | (24) | (24) | (24) | (24) | (24) | (24) |
| | **Volume (reps)** | 24-30 | 24-30 | 24-30 | 24-30 | 24-30 | 24-30 | 10-14 | 10-14 | 10-14 | 10-14 | 10-14 | 10-14 | 12 | 12 | 12 | 12 | 12 | 12 | 12 | 12 | 12 | 12 | 12 | 12 |
| **Aerobic training** | **Type** | Treadmill | | | | | | | | | | | | Treadmill | | | | | | | | | | | |
| | **Volume (min)** | 15 | 15 | 15 | 15 | 15 | 15 | 15 | 15 | 15 | 15 | 15 | 15 | 20 | 20 | 20 | 20 | 20 | 20 | 20 | 20 | 20 | 20 | 20 | 20 |
| | **Intensity** | ≤65% HRR | | | | | | | | | | | | 65-70% HRR | | | | | | | | | | | |
| | **Cool down** | 5 min of static and dynamic flexibility exercises | | | | | | | | | | | | 5 min of static and dynamic flexibility exercises | | | | | | | | | | | |

| | **Phases** | **Phase 2** | | | | | | **Phase 3** | | | | | | | | | | | | | | | | | |
|---|---|---|---|---|---|---|---|---|---|---|---|---|---|---|---|---|---|---|---|---|---|---|---|---|---|
| | **Weeks** | **9** | | | **10** | | | **11** | | | **12** | | | **13** | | | **14** | | | **15** | | | **16** | | |
| | **Session** | ***25*** | ***26*** | ***27*** | ***28*** | ***29*** | ***30*** | ***31*** | ***32*** | ***33*** | ***34*** | ***35*** | ***36*** | ***37*** | ***38*** | ***39*** | ***40*** | ***41*** | ***42*** | ***43*** | ***44*** | ***45*** | ***46*** | ***47*** | ***48*** |
| | **Warm-up** | 5 min of low intensity aerobic activity (50-65% HRR) on Treadmill | | | | | | | | | | | | | | | | | | | | | | | |
| | **Compensatory training *** | Core stability and stabiliser muscles (Level A-B) | | | | | | Core stability and stabiliser muscles (Level A-B-C) | | | | | | | | | | | | | | | | | |
| **Resis tance traini** | **Objective** | Metabolic stress / Mechanial tension and muscle damage | | | | | | Mechanical tension and muscle damage | | | | | | | | | | | | | | | | | |

| | | | | | | | | | | | | | | | | | | | | | | | | | |
|---|---|---|---|---|---|---|---|---|---|---|---|---|---|---|---|---|---|---|---|---|---|---|---|---|---|
| | **Type** | 6 main resistance exercises focused in major muscle groups | | | | | | 6 main resistance exercises focused in major muscle groups | | | | | | | | | | | | | | | | | |
| | **Sets** | 3 | 3 | 3 | 3 | 3 | 3 | 3 | 3 | 3 | 3 | 3 | 3 | 3 | 3 | 3 | 3 | 3 | 3 | 3 | 3 | 3 | 3 | 3 | 3 |
| | **Repetitions** | 12 | 12 | 12 | 12 | 12 | 12 | 10 | 10 | 10 | 10 | 10 | 10 | 8 | 8 | 8 | 8 | 8 | 8 | 6 | 6 | 6 | 6 | 6 | 6 |
| | **Intensity (LE)** | (24) | (24) | (24) | (24) | (24) | (24) | (20) | (20) | (20) | (20) | (20) | (20) | (16) | (16) | (16) | (16) | (16) | (16) | (12) | (12) | (12) | (10) | (10) | (10) |
| | **Volume (reps)** | 36 | 36 | 36 | 36 | 36 | 36 | 30 | 30 | 30 | 30 | 30 | 30 | 24 | 24 | 24 | 24 | 24 | 24 | 18 | 18 | 18 | 18 | 18 | 18 |
| **Aerobic training** | **Type** | Treadmill | | | | | | Treadmill | | | | | | | | | | | | | | | | | |
| | **Volume (min)** | 20 | 20 | 20 | 20 | 20 | 20 | 20 | 20 | 20 | 25 | 25 | 25 | 25 | 25 | 25 | 25 | 25 | 25 | 25 | 25 | 25 | 25 | 25 | 25 |
| | **Intensity** | 70-75% HRR | | | | | | 75-85% HRR | | | | | | | | | | | | | | | | | |
| | **Cool down** | 5 min of static and dynamic flexibility exercises | | | | | | 5 min of static and dynamic flexibility exercises | | | | | | | | | | | | | | | | | |

LE, level of effort (number of repetitions actually performed out of the maximum number of repetitions that could be performed with the current load). *Compensatory training: isometric exercises (20-30 s) and isokinetic movements (5-7 repetitions).

**Table S4.** Changes from week 16 to 1-year follow-up in sex hormone levels and ovarian function with per-protocol analysis.

| | **Bariatric surgery + usual care (n=24)** | | | **Bariatric surgery + exercise (n=18)** | | |
|---|---|---|---|---|---|---|
| | **Week 16 Mean (95% CI)** | **1-year Mean (95% CI)** | **Mean change (95% CI)** | **Week 16 Mean (95% CI)** | **1-year Mean (95% CI)** | **Mean change (95% CI)** |
| SHBG (nmol/L) | 96.0 (73.8 to 118.2) | 106.3 (83.5 to 129.1) | 10.4 (-14.6 to 35.3) | 106.9 (80.3 to 133.5) | 125.0 (97.2 to 152.8) | 18.1 (-12.7 to 48.9) |
| AMH (ng/mL) | 2.1 (1.0 to 3.3) | 2.1 (0.9 to 3.2) | -0.1 (-0.8 to 0.6) | 2.6 (1.2 to 4.0) | 2.7 (1.3 to 4.1) | 0.1 (-0.7 to 0.9) |
| FSH (mIU/mL) | 7.5 (5.9 to 9.2) | 7.6 (5.9 to 9.4) | 0.1 (-2.74 to 2.92) | 7.4 (5.4 to 9.4) | 7.1 (5.0 to 9.3) | -0.3 (-3.7 to 3.2) |
| LH (mIU/mL) | 6.8 (2.7 to 10.9) | 12.2 (7.8 to 16.5) | 5.3 (-1.9 to 12.6) | 7.5 (2.5 to 12.4) | 6.7 (1.3 to 12.1) | -0.8 (-9.6 to 8.1) |
| TSH (µIU/mL) | 1.8 (1.2 to 2.4) | 1.7 (1.0 to 2.3) | -0.1 (-0.8 to 0.5) | 1.3 (0.6 to 2.1) | 1.5 (0.7 to 2.3) | 0.2 (-0.6 to 1.0) |
| Free T4 (ng/dL) | 0.9 (0.8 to 0.9) | 0.8 (0.8 to 0.9) | -0.02 (-0.1 to 0.04) | 0.9 (0.8 to 0.9) | 0.9 (0.8 to 1.0) | 0.02 (-0.1 to 0.1) |
| Prolactin (ng/mL) | 11.8 (9.4 to 14.3) | 11.7 (9.2 to 14.2) | -0.1 (-2.9 to 2.6) | 11.1 (8.1 to 14.0) | 11.8 (8.7 to 14.9) | 0.7 (-2.7 to 4.1) |
| Estradiol (pmol/mL) | 445.0 (250.0 to 640.0) | 472.0 (268.0 to 675.0) | 26.5 (-266.0 to 319.0) | 405.0 (169.0 to 640.0) | 518.0 (266.0 to 770.0) | 113.4 (-247.0 to 474.0) |
| Progesterone (ng/mL) | 2.1 (0.6 to 3.7) | 3.1 (1.5 to 4.8) | 1.0 (-1.5 to 3.5) | 2.8 (0.9 to 4.7) | 1.6 (-0.4 to 3.7) | -1.2 (-4.3 to 1.9) |
| Testosterone (nmol/L) | 1.2 (0.9 to 1.5) | 1.1 (0.9 to 1.4) | -0.1 (-0.4 to 0.2) | 1.2 (0.9 to 1.6) | 1.2 (0.9 to 1.6) | -0.003 (-0.4 to 0.4) |
| Androstendione (ng/L) | 1.3 (0.8 to 1.7) | 1.3 (0.8 to 1.8) | 0.1 (-0.5 to 0.7) | 1.8 (1.2 to 2.4) | 1.8 (1.2 to 2.4) | 0.1 (-0.7 to 0.8) |
| FAI | 1.5 (0.7 to 2.3) | 1.1 (0.3 to 2.0) | -0.3 (-1.3 to 0.6) | 1.9 (0.9 to 2.8) | 1.8 (0.8 to 2.9) | -0.03 (-1.2 to 1.2) |
| DHEA-S (µg/dL) | 143.0 (111.3 to 174.0) | 126.0 (94.5 to 158.0) | -16.5 (-39.6 to 6.7) | 145.0 (108.4 to 183.0) | 140.0 (102.3 to 178.0) | -5.4 (-33.9 to 23.2) |
| Oocyte count (No) | 13.1 (10.0 to 16.2) | 11.3 (8.2 to 14.5) | -1.7 (-3.3 to 0.2) | 13.2 (9.6 to 16.9) | 13.1 (9.5 to 16.8) | -0.1 (-2.0 to 1.9) |
| Endometrial thickness (mm) | 6.1 (4.8 to 7.3) | 7.1 (5.8 to 8.4) | 1.0 (-1.1 to 3.1) | 6.6 (5.0 to 8.1) | 6.3 (4.6 to 7.9) | -0.3 (-2.9 to 2.3) |
| Right ovarian volumen ($cm^3$) | 6.6 (5.3 to 7.9) | 7.3 (5.9 to 8.6) | 0.7 (-1.0 to 2.3) | 6.2 (4.6 to 7.8) | 6.0 (4.3 to 7.7) | -0.2 (-2.2 to 1.9) |
| Left ovarian volumen ($cm^3$) | 4.9 (3.2 to 6.7) | 5.3 (3.6 to 7.1) | 0.4 (-1.5 to 2.3) | 6.1 (4.0 to 8.2) | 5.9 (3.7 to 8.0) | -0.2 (-2.5 to 2.1) |

| | | | | | | |
|---|---|---|---|---|---|---|
| Right ovary mean diameter (mm) | 23.8 (22.0 to 25.5) | 23.8 (22.0 to 25.6) | 0.02 (-2.2 to 2.3) | 22.6 (20.5 to 24.7) | 23.1 (20.8 to 25.3) | 0.5 (-2.3 to 3.3) |
| Left ovary mean diameter (mm) | 21.0 (18.9 to 23.1) | 20.9 (18.8 to 23.1) | -0.03 (-2.6 to 2.5) | 22.3 (19.8 to 24.9) | 22.0 (19.3 to 24.7) | -0.3 (-3.5 to 2.8) |
| UtA-PI | 3.4 (3.0 to 3.8) | 2.9 (2.5 to 3.4) | -0.5 (-1.1 to 0.1) | 3.6 (3.1 to 4.1) | 2.7 (2.1 to 3.2) | -0.9 (-1.7 to 0.2) |

SHBG, sex hormone-binding globulin; AMH, anti-müllerian hormone; FSH, follicle stimulating hormone; LH, luteinizing hormone; TSH, thyroid-stimulating hormone; FAI, free androgen index; DHEA-S, dehydroepiandrosterone sulfate; UtA-PI, uterine artery mean pulsatility index.

**Table S5.** Changes from week 16 to 1-year follow-up in anthropometric, cardiometabolic and inflammatory profile with per-protocol analysis.

| | **Bariatric surgery + usual care (n=24)** | | | **Bariatric surgery + exercise (n=18)** | | |
|---|---|---|---|---|---|---|
| | **Week 16 Mean (95% CI)** | **1-year Mean (95% CI)** | **Mean change (95% CI)** | **Week 16 Mean (95% CI)** | **1-year Mean (95% CI)** | **Mean change (95% CI)** |
| ***Anthropometric profile*** | | | | | | |
| Weight (kg) | 97.7 (91.1 to 104.3) | 78.8 (72.1 to 85.5) | -18.9 (-22.8 to 15.0) | 88.8 (80.9 to 96.7) | 72.8 (64.8 to 80.7) | -16.0 (-20.8 to 11.3) |
| Weight loss (%) | 22.0 (21.0 to 24.0) | 38.0 (36.0 to 41.0) | 15.0 (14.0 to 17.0) | 25.0 (23.0 to 28.0) | 39.0 (36.0 to 42.0) | 13.0 (11.0 to 16.0) |
| BMI (kg/m$^2$) | 37.0 (34.5 to 39.4) | 29.7 (27.2 to 32.2) | -7.3 (-8.7 to 5.8) | 33.8 (30.8 to 36.7) | 27.6 (24.7 to 30.6) | -6.2 (-8.0 to 4.3) |
| Body fat (%) | 46.3 (44.0 to 48.7) | 34.4 (32.0 to 36.9) | -11.9 (-14.3 to 9.5) | 42.5 (39.6 to 45.4) | 32.1 (29.1 to 35.1) | -10.4 (-13.3 to 7.4) |
| FFM (kg) | 51.8 (49.0 to 54.6) | 50.5 (47.7 to 53.4) | -1.3 (-3.1 to 0.5) | 51.2 (47.8 to 54.7) | 49.6 (46.1 to 53.1) | -1.7 (-3.8 to 0.5) |
| SMM (kg) | 28.4 (26.7 to 30.1) | 27.6 (25.9 to 29.3) | -0.9 (-1.94 to 0.2) | 28.0 (26.0 to 30.1) | 27.0 (24.9 to 29.1) | -1.0 (-2.34 to 0.3) |
| ***Cardiometabolic profile*** | | | | | | |
| Glucose (mg/dL) | 90.3 (85.9 to 94.7) | 83.0 (78.2 to 87.9) | -7.2 (-14.3 to 0.1) | 87.2 (81.9 to 92.5) | 82.4 (76.7 to 88.1) | -4.8 (-13.3 to 3.6) |
| Insulin (µUl/mL) | 8.6 (5.7 to 11.5) | 5.1 (2.0 to 8.2) | -3.5 (-7.5 to 0.4) | 6.6 (3.1 to 10.1) | 5.2 (1.5 to 8.9) | -1.4 (-6.1 to 3.3) |
| HOMA-IR index | 2.0 (1.2 to 2.8) | 1.1 (0.2 to 1.9) | -0.9 (-2.1 to 0.3) | 1.4 (0.5 to 2.4) | 1.1 (0.01 to 2.1) | -0.4 (-1.8 to 1.0) |
| Glycated hemoglobin (mg/dL) | 5.1 (4.9 to 5.3) | 5.0 (4.8 to 5.2) | -0.1 (-0.3 to 0.1) | 5.1 (4.9 to 5.3) | 5.1 (4.9 to 5.2) | -0.1 (-0.3 to 0.1) |
| Cortisol (µg/dL) | 9.1 (7.6 to 10.7) | 10.7 (9.0 to 12.4) | 1.6 (0.9 to 4.2) | 10.4 (8.6 to 12.3) | 11.3 (9.3 to 13.3) | 0.9 (-2.1 to 4.0) |
| HDL (mg/dL) | 44.8 (41.2 to 48.4) | 55.8 (51.9 to 59.7) | 11.0 (-6.4 to 15.6) | 42.4 (38.1 to 46.8) | 54.4 (49.8 to 58.9) | 11.9 (-6.5 to 17.4) |
| LDL (mg/dL) | 113.0 (98.6 to 127.0) | 104.5 (89.5 to 120.0) | -8.5 (-23.5 to 6.6) | 110.5 (93.4 to 128.0) | 92.1 (74.4 to 110.0) | -18.4 (-36.1 to 0.6) |
| Triglycerides (mg/dL) | 100.3 (86.2 to 114.3) | 67.5 (52.0 to 82.9) | -32.8 (-54.8 to 10.8) | 84.7 (67.7 to 101.7) | 65.8 (47.7 to 84.0) | -18.8 (-45.0 to 7.3) |
| Cholesterol (mg/dL) | 178.0 (162.0 to 193.0) | 173.0 (157.0 to 189.0) | -4.5 (-21.1 to 12.2) | 169.0 (151.0 to 188.0) | 160.0 (141.0 to 179.0) | -9.7 (-29.4 to 10.1) |

| | | | | | | |
|---|---|---|---|---|---|---|
| Aspartate transaminase (U/L) | 29.1 (25.1 to 33.0) | 25.1 (20.7 to 29.4) | -4.0 (-10.2 to 2.3) | 24.3 (19.5 to 29.2) | 22.6 (17.4 to 27.7) | -1.7 (-9.1 to 5.7) |
| Alanine transaminase (U/L) | 33.6 (26.3 to 41.0) | 28.3 (20.3 to 36.3) | -5.3 (-16.1 to 5.5) | 28.2 (19.3 to 37.1) | 26.4 (17.0 to 35.9) | -1.7 (-14.5 to 11.0) |
| Alkaline phosphatase (U/L) | 91.8 (81.3 to 102.3) | 87.8 (76.8 to 98.7) | -4.1 (-14.1 to 6.0) | 88.6 (76.3 to 101.3) | 87.2 (74.3 to 100.1) | -1.6 (-13.5 to 10.3) |
| Creatinine (mg/dL) | 0.6 (0.5 to 0.6) | 0.6 (0.6 to 0.6) | 0.004 (-0.03 to 0.04) | 0.6 (0.5 to 0.6) | 0.6 (0.5 to 0.6) | 0.02 (-0.02 to 0.1) |
| Glomerural filtration rate (mL/min) | 118.0 (114.0 to 122.0) | 118.0 (114.0 to 123.0) | 0.2 (-4.2 to 4.7) | 120.0 (115.0 to 124.0) | 118.0 (113.0 to 123.0) | -1.5 (-6.7 to 3.8) |
| Uric acid (mg/dL) | 5.2 (4.7 to 5.7) | 4.3 (3.8 to 4.8) | -0.9 (-1.6 to 0.3) | 4.6 (4.0 to 5.2) | 4.0 (3.4 to 4.7) | -0.6 (-1.4 to 0.1) |
| ***Inflammation*** | | | | | | |
| TNF-alpha (pg/mL) | 6.9 (6.1 to 7.8) | 2.7 (1.8 to 3.7) | -4.21 (-5.8 to 2.6) | 5.5 (4.4 to 6.5) | 3.3 (2.1 to 4.4) | -2.2 (-4.1 to 0.3) |
| Leptin (ng/mL) | 80.1 (64.0 to 96.2) | 58.0 (40.6 to 75.3) | -22.2 (-43.9 to 0.4) | 61.8 (42.5 to 81.2) | 45.4 (24.9 to 65.8) | -16.5 (-42.2 to 9.3) |
| C-reactive protein (mg/dL) | 0.6 (0.3 to 0.8) | 0.2 (-0.1 to 0.5) | -0.4 (-0.8 to 0.1) | 0.4 (0.03 to 0.7) | 0.2 (-0.2 to 0.6) | -0.2 (-0.7 to 0.3) |
| Pulse wave velocity (m/s) | 5.8 (5.5 to 6.1) | 5.4 (5.1 to 5.7) | -0.4 (-0.6 to 0.2) | 5.7 (5.3 to 6.0) | 5.5 (5.2 to 5.9) | -0.1 (-0.4 to 0.1) |
| ***Physical fitness*** | | | | | | |
| Handgrip strength (kg) | 28.6 (26.9 to 30.3) | 28.5 (26.9 to 30.1) | -0.1 (-6.0 to 5.8) | 28.2 (26.4 to 30.3) | 28.5 (24.4 to 29.2) | 0.3 (-0.4 to 1.0) |
| Chair stand test (repetitions) | 12.3 (11.3 to 13.4) | 13.7 (12.2 to 15.1) | 1.4 (-2.2 to 5.0) | 14.4 (12.6 to 16.3) | 16.2 (13.8 to 18.6) | 1.8 (-1.0 to 4.6) |
| Bruce test (s) | 471.0 (425.0 to 516.0) | 570.0 (535.0 to 604.0) | 99.0 (80.4 to 117.6) | 517.0 (434.0 to 600.0) | 570.0 (535.0 to 666.0) | 53.0 (-44.8 to 150.8) |
| Back scratch (cm) | -2.1 (-4.7 to 0.6) | -0.4 (-2.7 to 1.9) | 1.7 (-0.6 to 5.0) | -3.2 (-6.5 to 0.02) | -0.3 (-3.6 to 3.0) | 2.9 (-1.4 to 7.2) |

BMI, body mass index; FFM, fat free mass; SMM, skeletal muscle mass; HOMA-IR, homeostatic model assessment for insulin resistance; HDL, high-density lipoprotein; LDL, low-density lipoprotein; TNF-alpha, tumor necrosis factor alpha.

**Table S6.** Changes in sex hormone levels and ovarian function after 16-week of supervised exercise intervention and after a follow- up of 1-year post bariatric surgery with intention-to-treat analysis.

| End point | Bariatric surgery + usual care (n = 25) | Bariatric surgery + exercise (n = 21) | Mean difference between groups |
|---|---|---|---|
| SHBG (nmol/L) | | | |
| Baseline | 59.7 (37.9 to 81.6) | 47.4 (22.9 to 71.9) | - |
| Change at week 16 | 35.3 (10.9 to 59.8) | 65.9 (38.3 to 93.5) | 30.6 (0.1 to 61.1) |
| Change at 1-year | 44.9 (20.1 to 69.8) | 76.5 (47.7 to 105.3) | 31.5 (0.1 to 63.0) |
| AMH (ng/mL) | | | |
| Baseline | 2.8 (1.7 to 3.9) | 2.8 (1.6 to 4.1) | - |
| Change at week 16 | -0.7 (-1.3 to 0.1) | -0.5 (-1.2 to 0.2) | 0.2 (-0.6 to 1.0) |
| Change at 1-year | -0.8 (-1.4 to 0.1) | -0.3 (-1.1 to 0.4) | 0.4 (-0.4 to 1.3) |
| FSH (mIU/mL) | | | |
| Baseline | 7.3 (5.7 to 8.9) | 6.8 (4.9 to 8.6) | - |
| Change at week 16 | 0.2 (-2.5 to 3.0) | 0.4 (-2.8 to 3.5) | 0.1 (-3.3 to 3.6) |
| Change at 1-year | 0.5 (-2.3 to 3.3) | 0.7 (-2.5 to 4.0) | 0.2 (-3.3 to 3.7) |
| LH (mIU/mL) | | | |
| Baseline | 9.7 (5.5 to 13.8) | 7.0 (2.2 to 11.8) | - |
| Change at week 16 | -2.8 (-9.8 to 4.2) | 0.0 (-8.0 to 8.0) | 2.8 (-6.0 to 11.6) |
| Change at 1-year | 3.3 (-3.7 to 10.4) | -0.7 (-9.0 to 7.6) | -4.1 (-13.1 to 5.0) |
| TSH (µIU/mL) | | | |
| Baseline | 2.2 (1.6 to 2.8) | 1.8 (1.1 to 2.4) | - |
| Change at week 16 | -0.4 (-1.0 to 0.2) | -0.5 (-1.2 to 0.2) | -0.1 (-0.9 to 0.6) |
| Change at 1-year | -0.5 (-1.1 to 0.1) | -0.2 (-0.9 to 0.6) | 0.3 (-0.4 to 1.1) |
| Free T4 (ng/dL) | | | |
| Baseline | 0.89 (0.84 to 0.94) | 0.9 (0.9 to 1.0) | - |
| Change at week 16 | -0.02 (-0.1 to 0.1) | -0.1 (-0.01 to 0.1) | -0.1 (-0.1 to 0.04) |
| Change at 1-year | -0.04 (-0.1 to 0.02) | -0.0 (-0.01 to 0.1) | 0.0 (-0.1 to 0.1) |
| Prolactin (ng/mL) | | | |
| Baseline | 12.7 (10.3 to 15.1) | 11.9 (9.2 to 14.6) | - |
| Change at week 16 | -1.0 (-3.6 to 1.7) | -0.7 (-3.7 to 2.3) | 0.3 (-3.0 to 3.5) |
| Change at 1-year | -1.0 (-3.7 to 1.8) | -0.1 (-3.2 to 3.1) | 0.9 (-2.5 to 4.3) |
| Estradiol (pmol/L) | | | |
| Baseline | 339.0 (145.0 to 533.0) | 373.0 (157.0 to 589.0) | - |
| Change at week 16 | 105.7 (-180.0 to 391.0) | 45.1 (-276.0 to 366.0) | -60.6 (-416.3 to 295.2) |
| Change at 1-year | 126.8 (-163.0 to 416.0) | 111.1 (-223.0 to 445.0) | -15.7 (-381.5 to 350.0) |
| Progesterone (ng/mL) | | | |
| Baseline | 1.5 (-0.2 to 3.2) | 2.7 (0.8 to 4.7) | - |
| Change at week 16 | 0.7 (-1.9 to 3.2) | 0.5 (-2.5 to 3.4) | -0.2 (-3.4 to 3.0) |
| Change at 1-year | 1.5 (-1.1 to 4.1) | -1.2 (-4.2 to 1.9) | -2.7 (-6.0 to 0.6) |
| Testosterone (nmol/L) | | | |
| Baseline | 1.5 (1.2 to 1.8) | 1.5 (1.2 to 1.8) | - |
| Change at week 16 | -0.2 (-0.5 to 0.04) | -0.4 (-0.7 to 0.05) | -0.1 (-0.5 to 0.2) |
| Change at 1-year | -0.3 (0.6 to 0.001) | -0.4 (-0.8 to 0.1) | -0.1 (-0.5 to 0.3) |
| Androstendione (ng/mL) | | | |
| Baseline | 1.4 (0.9 to 1.9) | 1.8 (1.2 to 2.3) | - |
| Change at week 16 | -0.1 (-0.7 to 0.4) | -0.2 (-0.8 to 0.5) | -0.02 (-0.7 to 0.7) |
| Change at 1-year | -0.1 (-0.7 to 0.5) | -0.1 (-0.8 to 0.5) | -0.04 (-0.8 to 0.7) |

| | | | |
|---|---|---|---|
| FAI | | | |
| Baseline | 3.6 (2.8 to 4.4) | 3.8 (2.8 to 4.7) | - |
| Change at week 16 | -2.2 (-3.1 to 1.2) | -2.1 (-3.1 to 1.0) | 0.1 (-1.1 to 1.3) |
| Change at 1-year | -2.4 (-3.4 to 1.5) | -2.1 (-3.2 to 1.0) | 0.3 (-0.9 to 1.5) |
| DHEA-S (µg/dL) | | | |
| Baseline | 156.0 (124.3 to 187.0) | 156.0 (120.4 to 191.0) | - |
| Change at week 16 | -13.3 (-35.7 to 9.0) | -22.2 (-47.5 to 3.2) | -8.8 (-36.5 to 18.9) |
| Change at 1-year | -28.8 (-51.6 to 6.0) | -26.9 (-53.4 to 0.3) | 1.9 (-26.7 to 30.5) |
| Oocyte count (No) | | | |
| Baseline | 12.6 (9.5 to 15.7) | 12.1 (8.7 to 15.5) | - |
| Change at week 16 | 0.2 (-1.5 to 1.9) | 1.0 (-0.9 to 2.9) | 0.8 (-1.4 to 2.9) |
| Change at 1-year | -1.3 (-3.0 to 0.5) | 0.8 (-1.2 to 2.9) | 2.1 (-0.1 to 4.3) |
| Endometrial thickness (mm) | | | |
| Baseline | 7.9 (6.6 to 9.1) | 5.2 (3.7 to 6.6) | - |
| Change at week 16 | -1.7 (-3.8 to 0.4) | 1.0 (-1.4 to 3.3) | **2.7 (0.1 to 5.3)*** |
| Change at 1-year | -0.7 (-2.8 to 1.5) | 1.2 (-1.3 to 3.7) | 1.9 (-0.8 to 4.6) |
| Right ovarian volumen ($cm^3$) | | | |
| Baseline | 5.2 (3.9 to 6.5) | 6.0 (4.5 to 7.5) | - |
| Change at week 16 | 1.3 (-0.3 to 3.0) | 0.1 (-1.8 to 1.9) | -1.3 (-3.3 to 0.8) |
| Change at 1-year | 2.1 (-0.4 to 3.7) | -0.3 (-2.3 to 1.6) | **-2.4 (-4.5 to -0.3)*** |
| Left ovarian volumen ($cm^3$) | | | |
| Baseline | 5.2 (3.5 to 6.9) | 4.7 (2.8 to 6.6) | - |
| Change at week 16 | -0.2 (-2.0 to 1.7) | 1.2 (-0.9 to 3.2) | 1.3 (-1.0 to 3.6) |
| Change at 1-year | 0.5 (-1.4 to 2.3) | 1.1 (-1.1 to 3.3) | 0.6 (-1.8 to 3.0) |
| Right ovary mean diameter (mm) | | | |
| Baseline | 21.5 (19.8 to 23.3) | 22.2 (20.2 to 24.2) | - |
| Change at week 16 | 2.2 (-0.1 to 4.5) | 0.3 (-2.2 to 2.9) | -1.9 (-4.7 to 1.0) |
| Change at 1-year | 2.3 (-0.0 to 4.7) | 0.4 (-2.3 to 3.1) | -1.9 (-4.9 to 1.0) |
| Left ovary mean diameter (mm) | | | |
| Baseline | 21.0 (18.9 to 23.0) | 20.2 (17.9 to 22.5) | - |
| Change at week 16 | 0.1 (-2.5 to 2.6) | 2.0 (-0.9 to 4.8) | 1.9 (-1.2 to 5.0) |
| Change at 1-year | 0.3 (-2.3 to 2.9) | 1.9 (-1.1 to 4.9) | 1.6 (-1.7 to 4.8) |
| UtA-PI | | | |
| Baseline | 2.9 (2.5 to 3.4) | 3.9 (3.4 to 4.4) | - |
| Change at week 16 | 0.5 (-0.1 to 1.1) | -0.3 (-1.0 to 0.4) | **0.8 (-0.0 to 1.5)*** |
| Change at 1-year | -0.04 (-0.6 to 0.6) | -1.2 (-2.0 to 0.5) | **1.2 (-0.4 to 2.0)**** |

SHBG, sex hormone-binding globulin; AMH, anti-müllerian hormone; FSH, follicle stimulating hormone; LH, luteinizing hormone; TSH, thyroid-stimulating hormone; FAI, free androgen index; DHEA-S, dehydroepiandrosterone sulfate; UtA-PI, uterine artery mean pulsatility index; *, $p < 0.05$ time × group interaction; **, $p < 0.01$ time × group interaction.

**Table S8.** Changes from week 16 to 1-year follow-up in sex hormone levels and ovarian function with intention-to-treat analysis.

| | **Bariatric surgery + usual care (n = 25)** | | | **Bariatric surgery + exercise (n = 21)** | | |
|---|---|---|---|---|---|---|
| | **Week 16 Mean (95% CI)** | **1-year Mean (95% CI)** | **Mean change (95% CI)** | **Week 16 Mean (95% CI)** | **1-year Mean (95% CI)** | **Mean change (95% CI)** |
| SHBG (nmol/L) | 95.1 (73.5 to 116.6) | 104.7 (82.8 to 126.5) | 9.6 (-14.8 to 34.0) | 113.3 (88.8 to 137.8) | 123.9 (98.5 to 149.4) | 10.6 (-17.7 to 38.9) |
| AMH (ng/mL) | 2.1 (1.0 to 3.2) | 2.0 (0.9 to 3.1) | -0.1 (-0.7 to 0.6) | 2.4 (1.1 to 3.6) | 2.5 (1.2 to 3.8) | 0.2 (-0.6 to 0.9) |
| FSH (mIU/mL) | 7.5 (5.9 to 9.1) | 7.8 (6.2 to 9.5) | 0.3 (-2.5 to 3.1) | 7.1 (5.3 to 9.0) | 7.5 (5.5 to 9.4) | 0.4 (-2.8 to 3.6) |
| LH (mIU/mL) | 6.9 (2.8 to 10.9) | 13.0 (8.8 to 17.1) | 6.1 (-0.9 to 13.1) | 7.0 (2.4 to 11.6) | 6.3 (1.4 to 11.2) | -0.7 (-8.9 to 7.4) |
| TSH (µIU/mL) | 1.8 (1.2 to 2.4) | 1.7 (1.1 to 2.3) | -0.1 (-0.8 to 0.5) | 1.3 (0.6 to 1.9) | 1.6 (0.9 to 2.3) | 0.3 (-0.4 to 1.0) |
| Free T4 (ng/dL) | 0.9 (0.8 to 0.9) | 0.8 (0.8 to 0.9) | -0.03 (-0.1 to 0.04) | 0.9 (0.8 to 0.9) | 0.9 (0.8 to 1.0) | 0.02 (-0.1 to 0.1) |
| Prolactin (ng/mL) | 11.7 (9.3 to 14.1) | 11.7 (9.3 to 14.1) | -0.01 (-2.7 to 2.7) | 11.2 (8.5 to 13.9) | 11.9 (9.1 to 14.7) | 0.6 (-2.5 to 3.7) |
| Estradiol (pmol/mL) | 445.0 (255.0 to 634.0) | 466.0 (272.0 to 659.0) | 21.1 (-264.0 to 306.0) | 418.0 (202.0 to 634.0) | 484.0 (255.0 to 713.0) | 65.9 (-265.0 to 397.0) |
| Progesterone (ng/mL) | 2.1 (0.5 to 3.8) | 3.0 (1.3 to 4.7) | 0.9 (-1.7 to 3.4) | 3.2 (1.4 to 5.1) | 1.6 (-0.4 to 3.5) | -1.6 (-4.6 to 1.3) |
| Testosterone (nmol/L) | 1.2 (0.9 to 1.5) | 1.2 (0.9 to 1.5) | -0.1 (-0.3 to 0.2) | 1.1 (0.8 to 1.5) | 1.1 (0.8 to 1.4) | -0.03 (-0.4 to 0.3) |
| Androstenedione (ng/L) | 1.3 (0.8 to 1.7) | 1.3 (0.9 to 1.8) | 0.1 (-0.5 to 0.7) | 1.6 (1.1 to 2.1) | 1.7 (1.1 to 2.2) | 0.1 (-0.6 to 0.7) |
| FAI | 1.5 (0.7 to 2.3) | 1.2 (0.4 to 2.0) | -0.3 (-1.2 to 0.7) | 1.7 (0.8 to 2.6) | 1.6 (0.7 to 2.6) | -0.1 (-1.2 to 1.0) |
| DHEA-S (µg/dL) | 142.0 (112.2 to 174.0) | 127.0 (95.5 to 158.0) | -15.5 (-37.8 to 6.9) | 134.0 (98.2 to 169.0) | 129.0 (93.0 to 165.0) | -4.7 (-30.6 to 21.2) |
| Oocyte count (No) | 12.8 (9.7 to 15.9) | 11.3 (8.2 to 14.4) | -1.5 (-3.2 to 0.2) | 13.1 (9.7 to 16.5) | 13.0 (9.5 to 16.4) | -0.1 (-2.1 to 1.8) |
| Endometrial thickness (mm) | 6.2 (4.9 to 7.4) | 7.2 (5.9 to 8.5) | 1.0 (-1.1 to 3.2) | 6.1 (4.7 to 7.6) | 6.4 (4.8 to 7.9) | 0.2 (2.2 to 2.7) |
| Right ovarian volume ($cm^3$) | 6.5 (-7.8 to 5.2) | 7.3 (5.9 to 8.6) | 0.7 (-0.9 to 2.4) | 6.1 (4.6 to 7.5) | 5.7 (4.1 to 7.2) | -0.4 (-2.3 to 1.5) |
| Left ovarian volume ($cm^3$) | 5.0 (3.3 to 6.7) | 5.6 (3.9 to 7.3) | 0.6 (-1.2 to 2.5) | 5.9 (4.0 to 7.7) | 5.8 (3.8 to 7.7) | -0.1 (-2.2 to 2.0) |

| | | | | | | |
|---|---|---|---|---|---|---|
| Right ovary mean diameter (mm) | 23.7 (22.0 to 25.5) | 23.9 (22.1 to 25.6) | 0.1 (-2.1 to 2.4) | 22.5 (20.6 to 24.4) | 22.6 (20.5 to 24.6) | 0.1 (-2.6 to 2.7) |
| Left ovary mean diameter (mm) | 21.1 (19.0 to 23.1) | 21.3 (19.2 to 23.4) | 0.2 (-2.3 to 2.8) | 22.2 (20.0 to 24.4) | 22.1 (19.7 to 24.5) | -0.1 (-3.1 to 2.8) |
| UtA-PI | 3.4 (3.0 to 3.8) | 2.9 (2.5 to 3.3) | -0.5 (-1.1 to 0.1) | 3.6 (3.1 to 4.1) | 2.7 (2.2 to 3.2) | -0.9 (-1.6 to 0.2) |

SHBG, sex hormone-binding globulin; AMH, anti-müllerian hormone; FSH, follicle stimulating hormone; LH, luteinizing hormone; TSH, thyroid-stimulating hormone; FAI, free androgen index; DHEA-S, dehydroepiandrosterone sulfate; UtA-PI, uterine artery mean pulsatility index.

**Table S7.** Changes in anthropometric, cardiometabolic and inflammatory profile after 16-week of supervised exercise intervention and after a follow up of 1-year post bariatric surgery.

| End point | Bariatric surgery + usual care (n = 25) | Bariatric surgery + exercise (n = 21) | Mean difference between groups |
|---|---|---|---|
| ***Anthropometric profile*** | | | |
| Weight (kg) | | | |
| Baseline | 125.8 (119.2 to 132.3) | 116.1 (108.6 to 123.5) | - |
| Change at week 16 | -28.4 (-32.2 to -24.7) | -29.5 (-33.9 to -25.2) | -1.1 (-5.9 to 3.6) |
| Change at 1-year | -47.3 (-51.3 to -43.3) | -43.9 (-48.5 to -39.3) | 3.4 (-1.6 to 8.4) |
| Weight loss (%) | | | |
| Baseline | - | - | - |
| Change at week 16 | 23.0 (21.0 to 24.0) | 26.0 (24.0 to 29.0) | 3.0 (0.5 to 5.5) |
| Change at 1-year | 38.0 (36.0 to 41.0) | 38.0 (36.0 to 41.0) | 0.2 (0.02 to 0.03) |
| BMI (kg/m$^2$) | | | |
| Baseline | 47.4 (44.4 to 50.4) | 44.5 (41.8 to 47.2) | - |
| Change at week 16 | -10.7 (-12.1 to -9.2) | -11.4 (-13.1 to -9.7) | -0.7 (-2.5 to 1.3) |
| Change at 1-year | -17.9 (-19.4 to -16.4) | -16.9 (-18.7 to -15.1) | 1.0 (-0.9 to 2.9) |
| Body fat (%) | | | |
| Baseline | 53.2 (51.0 to 55.5) | 51.9 (49.3 to 54.5) | - |
| Change at week 16 | -6.9 (-9.3 to -4.6) | -10.0 (-12.8 to -7.3) | -3.1 (-6.0 to -0.1) |
| Change at 1-year | -18.8 (-21.3 to -16.4) | -19.4 (-22.3 to -16.5) | -0.6 (-3.7 to 2.5) |
| FFM (kg) | | | |
| Baseline | 58.6 (55.7 to 61.5) | 56.0 (52.6 to 59.3) | - |
| Change at week 16 | -6.9 (-8.6 to -5.3) | -5.8 (-7.8 to -3.9) | 1.1 (-1.0 to 3.2) |
| Change at 1-year | -8.2 (-9.9 to -6.4) | -7.3 (-9.3 to -5.3) | 0.9 (-1.3 to 3.1) |
| SMM (kg) | | | |
| Baseline | 33.1 (31.4 to 34.8) | 31.4 (29.4 to 33.4) | - |
| Change at week 16 | -4.8 (-5.8 to 3.8) | -4.0 (-5.2 to 2.9) | 0.7 (-0.6 to 2.0) |
| Change at 1-year | -5.6 (-6.7 to 4.5) | -4.9 (-6.2 to 3.7) | 0.7 (-0.7 to 2.0) |
| ***Cardiometabolic profile*** | | | |
| Glucose (Mg/dL) | | | |
| Baseline | 97.0 (92.7 to 101.4) | 98.0 (93.0 to 102.9) | - |
| Change at week 16 | -6.9 (-13.6 to -0.3) | -10.9 (-18.5 to -3.6) | -4.0 (-12.3 to 4.2) |
| Change at 1-year | -14.2 (-21.2 to -7.1) | -15.2 (-23.0 to -7.3) | -1.0 (-9.6 to 7.7) |
| Insulin (µUl/mL) | | | |
| Baseline | 17.6 (14.8 to 20.5) | 13.4 (10.1 to 16.6) | - |
| Change at week 16 | -9.2 (-12.8 to -5.6) | -7.1 (-11.2 to -2.9) | 2.1 (-2.5 to 6.7) |
| Change at 1-year | -12.7 (-16.6 to -8.9) | -8.3 (-12.6 to -4.0) | 4.4 (-0.4 to 9.2) |
| HOMA-IR | | | |
| Baseline | 4.4 (3.6 to 5.2) | 3.4 (2.5 to 4.3 | - |
| Change at week 16 | -2.5 (-3.6 to -1.4) | -2.0 (-3.3 to -0.8) | 0.4 ( -0.9 to 1.8) |
| Change at 1-year | -3.4 (-4.6 to -2.6) | -2.4 (-3.7 to -1.1) | 1.0 (-0.4 to 2.4) |
| Glycated hemoglobin (Mg/dL) | | | |
| Baseline | 5.3 (5.2 to 5.5) | 5.4 (5.2 to 5.6) | - |
| Change at week 16 | -0.2 (-0.4 to -0.1) | -0.3 (-0.5 to -0.1) | -0.1 (-0.3 to 0.1) |
| Change at 1-year | -0.4 (-0.5 to -0.2) | -0.4 (-0.6 to -0.2) | -0.0 (-0.2 to 0.2) |
| Cortisol (µg/dL) | | | |
| Baseline | 9.4 (7.9 to 10.9) | 10.2 (8.5 to 11.9 | - |
| Change at week 16 | -0.3 (-2.7 to 2.1) | -0.1 (-2.8 to 2.6) | 0.2 (-2.7 to 3.2) |

| | | | |
|---|---|---|---|
| Change at 1-year | 1.3 (-1.2 to 3.8) | 0.8 (-2.0 to 3.6) | -0.4 (-3.5 to 2.6) |
| HDL (Mg/dL) | | | |
| Baseline | 53.4 (49.7 to 57.1) | 51.9 (47.7 to 56.1) | - |
| Change at week 16 | -8.7 (-13.4 to 4.1) | -8.8 (-14.0 to 3.5) | -0.0 (-5.8 to 5.7) |
| Change at 1-year | 2.2 (-2.7 to 7.1) | 3.8 (-1.7 to 9.3) | 1.6 (-4.5 to 7.6) |
| LDL (Mg/dL) | | | |
| Baseline | 125.0 (111.2 to 140.0) | 115.0 (98.8 to 131.0) | - |
| Change at week 16 | -14.2 (-28.0 to 0.4) | -5.4 (-21.2 to 10.3) | 8.8 (-8.5 to 26.1) |
| Change at 1-year | -22.7 (-37.3 to 8.1) | -24.0 (-40.4 to 7.5) | -1.3 (-19.5 to 17.0) |
| Triglycerides (Mg/dL) | | | |
| Baseline | 118.8 (104.8 to 132.7) | 114.1 (98.2 to 129.9) | - |
| Change at week 16 | -19.5 (-40.0 to 1.0) | -28.7 (-52.0 to 5.3) | -9.2 (-34.6 to 16.2) |
| Change at 1-year | -52.4 (-74.0 to 30.7) | -47.9 (-72.1 to 23.6) | 4.5 (-22.1 to 31.1) |
| Cholesterol (Mg/dL) | | | |
| Baseline | 203.0 (187.0 to 218.0) | 190.0 (172.0 to 207.0) | - |
| Change at week 16 | -27.3 (-42.7 to 11.8) | -20.3 (-37.9 to 2.6) | 7.0 (-12.2 to 26.2) |
| Change at 1-year | -31.8 (-48.2 to 15.4) | -30.8 (-49.3 to 12.4) | 1.0 (-19.2 to 21.1) |
| Aspartate transaminase (U/L) | | | |
| Baseline | 24.2 (20.2 to 28.1) | 22.9 (18.4 to 27.4) | - |
| Change at week 16 | 4.9 (-1.0 to 10.7) | 1.2 (-5.4 to 7.9) | -3.6 (-10.9 to 3.6) |
| Change at 1-year | 0.9 (-5.3 to 7.1) | 0.8 (-6.1 to 7.7) | -0.1 (-7.7 to 7.5) |
| Alanine transaminase (U/L) | | | |
| Baseline | 28.4 (21.2 to 35.6) | 25.5 (17.3 to 33.7) | - |
| Change at week 16 | 5.4 (-4.6 to 15.5) | 1.7 (-9.8 to 13.2) | -3.7 (-16.2 to 8.8) |
| Change at 1-year | 0.2 (-10.5 to 10.9) | 1.9 (-10.1 to 13.9) | 1.7 (-11.4 to 14.8) |
| Alkaline phosphatase (U/L) | | | |
| Baseline | 83.4 (72.8 to 94.0) | 83.6 (71.6 to 95.5) | - |
| Change at week 16 | 7.1 (-2.3 to 16.5) | 4.6 (-6.1 to 15.4) | -2.5 (-14.2 to 9.2) |
| Change at 1-year | 3.1 (-6.9 to 13.0) | 5.6 (-5.7 to 16.8) | 2.5 (-9.8 to 14.8) |
| Creatinine (Mg/dL) | | | |
| Baseline | 0.7 (0.6 to 0.7) | 0.7 (0.6 to 0.7) | - |
| Change at week 16 | -0.1 (-0.1 to 0.02) | -0.1 (-0.1 to 0.03) | -0.0 (-0.1 to 0.04) |
| Change at 1-year | -0.1 (-0.1 to 0.01) | -0.1 (-0.1 to 0.01) | 0.0 (-0.1 to 0.1) |
| Glomerural filtration rate (mL/min) | | | |
| Baseline | 113.0 (109.0 to 117.0) | 114.0 (109.0 to 118.0) | - |
| Change at week 16 | 5.3 (-1.2 to 9.5) | 5.5 (-0.8 to 10.2) | 0.2 (-4.9 to 5.3) |
| Change at 1-year | 5.6 (-1.2 to 9.9) | 3.9 (-1.0 to 8.8)) | -1.7 (-7.0 to 3.7) |
| Uric acid (mg/dL) | | | |
| Baseline | 5.8 (5.3 to 6.3) | 5.5 (4.9 to 6.0) | - |
| Change at week 16 | -0.6 (-1.2 to 0.001) | -0.9 (-1.5 to 0.2) | -0.3 (-1.0 to 0.5) |
| Change at 1-year | -1.5 (-2.1 to 0.9) | -1.4 (-2.1 to 0.7) | 0.1 (-0.7 to 0.9) |
| ***Inflammation*** | | | |
| TNF-alpha (pg/mL) | | | |
| Baseline | 7.7 (6.8 to 8.6) | 7.9 (6.9 to 8.9) | - |
| Change at week 16 | -0.8 (-2.3 to 0.7) | -2.7 (-4.4 to 1.0) | **-1.9 (-3.8 to 0.0)*** |
| Change at 1-year | -5.0 (-6.6 to 3.4) | -4.6 (-6.4 to 2.9) | 0.4 (-1.6 to 2.3) |
| Leptin (ng/mL) | | | |

| | | | |
|---|---|---|---|
| Baseline | 194.3 (177.6 to 211.1) | 182.9 (164.1 to 201.6) | - |
| Change at week 16 | -115.8 (-137.1 to 94.5) | -123.7 (-147.7 to 99.8) | -7.9 (-34.4 to 18.6) |
| Change at 1-year | -138.2 (-160.8 to 115.6) | -137.0 (-162.2 to 112.1) | 1.2 (-26.6 to 29.0) |
| C-reactive protein (mg/dL) | | | |
| Baseline | 1.3 (1.0 to 1.6) | 0.8 (0.5 to 1.6) | - |
| Change at week 16 | 0.8 (0.4 to 1.2) | 0.5 (0.02 to 0.9) | 0.3 (-0.2 to 0.8) |
| Change at 1-year | 1.1 (0.7 to 1.6) | 0.6 (0.2 to 1.1) | 0.5 (-0.03 to 1.0) |
| Pulse wave velocity (m/s) | | | |
| Baseline | 6.0 (5.7 to 6.2) | 6.1 (5.8 to 6.4) | - |
| Change at week 16 | -0.2 (-0.4 to 0.04) | -0.4 (-0.6 to 0.1) | -0.2 (-0.5 to 0.1) |
| Change at 1-year | -0.6 (-0.8 to 0.4) | -0.5 (-0.8 to 0.2) | 0.1 (-0.2 to 0.4) |
| ***Physical fitness*** | | | |
| Handgrip strength (kg) | | | |
| Baseline | 30.4 (28.7 to 32.1) | 29.2 (27.1 to 31.2) | - |
| Change at week 16 | 1.8 (-0.1 to 3.7) | 1.6 (-0.8 to 4.0) | -0.2 (-2.8 to 2.4) |
| Change at 1-year | 1.9 (-0.1 to 3.9) | 3.0 (0.2 to 5.8) | 1.1 (-1.8 to 4.0) |
| Chair stand test (repetitions) | | | |
| Baseline | 10.4 (9.5 to 11.3) | 11.9 (10.3 to 13.4) | - |
| Change at week 16 | 1.9 (0.8 to 4.0) | 2.4 (0.4 to 4.4) | 0.5 (-1.7 to 2.7) |
| Change at 1-year | 3.3 (1.8 to 4.7) | 3.9 (1.4 to 6.4) | 0.6 (-1.9 to 3.1) |
| Bruce test (s) | | | |
| Baseline | 324.0 (274.0 to 373.0) | 372.0 (314.0 to 430.0) | - |
| Change at week 16 | 147.0 (92.6 to 201.4) | 151.0 (67.4 to 234.06) | 4.0 (-80.9 to 88.9) |
| Change at 1-year | 246.0 (195.9 to 296.1) | 386.0 (254.3 to 517.7) | 140.0 (17.8 to 262.2) |
| Back scratch (cm) | | | |
| Baseline | -12.5 (-15.5 to -9.5) | -11.4 (-14.6 to -8.1) | - |
| Change at week 16 | 10.0 (6.8 to 13.2) | 8.3 (4.5 to 12.1) | -1.7 (-5.9 to 2.5) |
| Change at 1-year | 8.3 (5.1 to 11.5) | 11.2 (7.2 to 15.2) | 2.9 (-1.4 to 7.2) |

BMI, body mass index; FFM, fat free mass; SMM, skeletal muscle mass; HOMA-IR, homeostatic model assessment for insulin resistance; HDL, high-density lipoprotein; LDL, low-density lipoprotein; TNF-alpha, tumor necrosis factor alpha; *, $p < 0.05$ time × group interaction.

**Table S9.** Changes from week 16 to 1-year follow-up in anthropometric, cardiometabolic and inflammatory profile with intention-to-treat analysis.

| | **Bariatric surgery + usual care (n = 25)** | | | **Bariatric surgery + exercise (n = 21)** | | |
|---|---|---|---|---|---|---|
| | **Week 16 Mean (95% CI)** | **1-year Mean (95% CI)** | **Mean change (95% CI)** | **Week 16 Mean (95% CI)** | **1-year Mean (95% CI)** | **Mean change (95% CI)** |
| ***Anthropometric profile*** | | | | | | |
| Weight (kg) | 97.3 (90.8 to 103.9) | 78.4 (71.8 to 85.1) | -18.9 (-22.9 to 14.9) | 86.5 (79.0 to 94.0) | 72.2 (64.6 to 79.7) | -14.4 (-18.9 to 9.8) |
| Weight loss (%) | 23.0 (21.0 to 24.0) | 26.0 (24.0 to 29.0) | 3.0 (0.2 to 6.0) | 38.0 (36.0 to 41.0) | 38.0 (36.0 to 41.0) | 0.02 (0.02 to 0.03) |
| BMI (kg/m$^2$) | 36.7 (34.3 to 39.1) | 29.4 (27.0 to 31.9) | -7.3 (-8.8 to 5.8) | 33.1 (30.4 to 35.9) | 27.6 (24.9 to 30.4) | -5.5 (-7.2 to 3.8) |
| Body fat (%) | 46.3 (44.1 to 48.5) | 34.4 (32.0 to 36.8) | -11.9 (-14.4 to 9.4) | 41.9 (39.3 to 44.5) | 32.5 (29.8 to 35.2) | -9.4 (-12.2 to 6.6) |
| FFM (kg) | 51.7 (48.8 to 54.5) | 50.4 (47.5 to 53.3) | -1.3 (-3.0 to 0.5) | 50.2 (46.8 to 53.5) | 48.7 (45.3 to 52.1) | -1.5 (-3.4 to 0.5) |
| SMM (kg) | 28.3 (26.6 to 30.0) | 27.5 (25.7 to 29.2) | -0.8 (-1.9 to 0.2) | 27.4 (25.4 to 29.4) | 26.5 (24.5 to 28.5) | -0.9 (-2.1 to 0.3) |
| ***Cardiometabolic profile*** | | | | | | |
| Glucose (mg/dL) | 90.1 (85.7 to 94.5) | 82.9 (78.1 to 87.7) | -7.2 (-14.3 to 0.2) | 87.0 (82.0 to 92.0) | 82.8 (77.6 to 88.1) | -4.2 (-12.0 to 3.6) |
| Insulin (µUl/mL) | 8.4 (5.6 to 11.3) | 4.9 (1.8 to 8.0) | -3.5 (-7.4 to 0.3) | 6.3 (3.1 to 9.5) | 5.1 (1.7 to 8.5) | -1.2 (-5.5 to 3.0) |
| HOMA-IR index | 1.9 (1.1 to 2.7) | 1.0 (0.1 to 1.9) | -0.9 (-2.1 to 0.2) | 1.4 (0.5 to 2.3) | 1.0 (0.1 to 2.0) | -0.3 (-1.6 to 0.9) |
| Glycated hemoglobin (mg/dL) | 5.1 (4.9 to 5.3) | 5.0 (4.8 to 5.2) | -0.1 (-0.3 to 0.1) | 5.1 (4.9 to 5.3) | 5.1 (4.9 to 5.2) | -0.03 (-0.2 to 0.2) |
| Cortisol (µg/dL) | 9.1 (7.6 to 10.6) | 10.7 (9.0 to 12.4) | 1.6 (-0.9 to 4.1) | 10.1 (8.4 to 11.9) | 11.1 (9.2 to 12.9) | 0.9 (-1.8 to 3.7) |
| HDL (mg/dL) | 44.7 (41.0 to 48.3) | 55.6 (51.6 to 59.6) | 10.9 (-6.0 to 15.9) | 43.2 (39.0 to 47.3) | 55.7 (51.3 to 60.1) | 12.5 (-7.10 to 18.0) |
| LDL (mg/dL) | 111.0 (97.0 to 126.0) | 103.0 (87.8 to 118.0) | -8.5 (-23.1 to 6.1) | 110.0 (93.4 to 126.0) | 91.0 (74.4 to 108.0) | -18.5 (-34.7 to 2.4) |
| Triglycerides (mg/dL) | 99.3 (85.3 to 113.2) | 66.4 (51.1 to 81.7) | -32.9 (-54.5 to 11.2) | 85.4 (69.5 to 101.2) | 66.2 (49.4 to 83.0) | -19.2 (-43.3 to 4.9) |
| Cholesterol (mg/dL) | 175.0 (160.0 to 191.0) | 171.0 (155.0 to 187.0) | -4.5 (-20.9 to 11.9) | 169.0 (152.0 to 187.0) | 159.0 (141.0 to 177.0) | -10.6 (-28.7 to 7.5) |

| | | | | | | |
|---|---|---|---|---|---|---|
| Aspartate transaminase (U/L) | 29.0 (25.1 to 32.9) | 25.1 (20.7 to 29.4) | -3.9 (-10.1 to 2.2) | 24.1 (19.7 to 28.6) | 23.7 (19.0 to 28.4) | -0.4 (-7.3 to 6.4) |
| Alanine transaminase (U/L) | 33.8 (26.7 to 41.0) | 28.6 (20.8 to 36.5) | -5.2 (-15.9 to 5.5) | 27.2 (19.1 to 35.4) | 27.4 (18.8 to 36.0) | 0.1 (-11.7 to 12.0) |
| Alkaline phosphatase (U/L) | 90.5 (80.0 to 101.1) | 86.5 (75.5 to 97.5) | -4.1 (-14.1 to 5.9) | 88.2 (76.2 to 100.2) | 89.1 (76.8 to 101.4) | 0.9 (-10.1 to 11.9) |
| Creatinine (mg/dL) | 0.6 (0.5 to 0.6) | 0.6 (0.5 to 0.6) | 0.01 (-0.03 to 0.05) | 0.6 (0.5 to 0.6) | 0.6 (0.5 to 0.6) | 0.02 (-0.02 to 0.1) |
| Glomerural filtration rate (mL/min) | 119.0 (115.0 to 122.0) | 119.0 (115.0 to 123.0) | 0.2 (-4.1 to 4.6) | 119.0 (115.0 to 123.0) | 117.0 (113.0 to 122.0) | -1.6 (-6.4 to 3.2) |
| Uric acid (mg/dL) | 5.2 (4.7 to 5.7) | 4.3 (3.8 to 4.8) | -0.9 (-1.6 to 0.3) | 4.6 (4.0 to 5.2) | 4.0 (3.5 to 4.6) | -0.6 (-1.3 to 0.1) |
| ***Inflammation*** | | | | | | |
| TNF-alpha (pg/mL) | 6.9 (6.0 to 7.8) | 2.7 (1.7 to 3.7) | -4.2 (-5.8 to 2.6) | 5.2 (4.2 to 6.2) | 3.3 (2.2 to 4.4) | -1.9 (-3.7 to 0.2) |
| Leptin (ng/mL) | 78.5 (62.0 to 95.0) | 56.1 (38.3 to 73.9) | -22.4 (-44.6 to 0.2) | 59.1 (40.4 to 77.9) | 45.9 (26.2 to 65.5) | -13.3 (-37.9 to 11.3) |
| C-reactive protein (mg/dL) | 0.5 (0.3 to 0.8) | 0.2 (-0.1 to 0.5) | -0.4 (-0.8 to 0.1) | 0.4 (0.03 to 0.7) | 0.2 (-0.1 to 0.5) | -0.2 (-0.6 to 0.3) |
| Pulse wave velocity (m/s) | 5.8 (5.5 to 6.0) | 5.4 (5.1 to 5.6) | -0.4 (-0.6 to 0.2) | 5.7 (5.4 to 6.0) | 5.6 (5.3 to 5.9) | -0.1 (-0.4 to 0.1) |
| ***Physical fitness*** | | | | | | |
| Handgrip strength (kg) | 28.6 (26.9 to 31.2) | 28.5 (26.9 to 30.1) | -0.1 (-6.0 to 5.8) | 27.6 (25.8 to 31.2) | 26.2 (23.9 to 28.5) | -1.4 (-4.7 to 1.9) |
| Chair stand test (repetitions) | 11.9 (10.3 to 13.3) | 13.7 (12.2 to 15.1) | 1.8 (0.1 to 3.5) | 14.3 (12.6 to 15.9) | 15.8 (13.6 to 18.0) | 1.5 (-1.0 to 4.0) |
| Bruce test (s) | 471.0 (425.0 to 516.0) | 570.0 (535.0 to 604.0) | 99.0 (53.3 to 114.7) | 523.0 (448.0 to 597.0) | 537.0 (423.0 to 651.0) | 14.0 (-111.7 to 134.7) |
| Back scratch (cm) | -2.1 (-4.7 to 0.6) | -0.4 (-2.7 to 1.9) | 1.7 (-1.1 to 4.5) | -3.1 (-5.9 to -0.1) | -0.2 (-3.1 to 2.8) | 2.9 (-0.9 to 5.7) |

BMI, body mass index; FFM, fat free mass; SMM, skeletal muscle mass; HOMA-IR, homeostatic model assessment for insulin resistance; HDL, high-density lipoprotein; LDL, low-density lipoprotein; TNF-alpha, tumor necrosis factor alpha.

**Table supplementary S10.** Changes in the food frequency questionnaire after 16-week of supervised exercise intervention and after 1-year post bariatric surgery with per-protocol analysis.

| End point | Bariatric surgery + usual care (n=24) | Bariatric surgery + exercise (n=18) | Mean difference between groups |
|---|---|---|---|
| Dairy products (servings/day) | | | |
| Baseline | 2.1 (1.5 to 2.7) | 2.4 (1.7 to 3.1) | - |
| Change at week 16 | -0.4 (-1.4 to 0.6) | 0.0 (-1.2 to 1.2) | 0.4 (-1.2 to 2.0) |
| Change at 1-year | 0.2 (-0.7 to 1.1) | 0.2 (-0.8 to 1.2) | 0.0 (-1.4 to 1.4) |
| Eggs (servings/day) | | | |
| Baseline | 2.4 (1.7 to 3.0) | 2.4 (1.7 to 3.2) | - |
| Change at week 16 | -0.1 (-1.0 to 0.8) | 0.2 (-0.8 to 1.2) | 0.3 (-1.1 to 1.7) |
| Change at 1-year | 11.2 (8.0 to 14.4) | 10.9 (7.9 to 13.8) | -0.3 (-4.7 to 4.1) |
| Meat products (servings/day) | | | |
| Baseline | 17.1 (13.2 to 21.0) | 16.7 (11.7 to 21.8) | - |
| Change at week 16 | -3.5 (-8.6 to 1.6) | -3.4 (-9.2 to 2.4) | 0.1 (-7.5 to 7.8) |
| Change at 1-year | -13.0 (-17.1 to 8.9) | -12.6 (-17.8 to 7.4) | 0.4 (-17.7 to 18.5) |
| Fish or seafood (servings/day) | | | |
| Baseline | 6.2 (4.1 to 8.3) | 10.1 (5.8 to 14.3) | - |
| Change at week 16 | 1.4 (-1.5 to 5.7) | 0.1 (-4.9 to 5.2) | -1.3 (-7.5 to 4.9) |
| Change at 1-year | -4.6 (-6.8 to 2.4) | -8.2 (-12.6 to 3.8) | -3.6 to (-12.9 to 5.8) |
| Vegetables (servings/day) | | | |
| Baseline | 4.5 (3.2 to 5.7) | 4.2 (3.2 to 5.2) | - |
| Change at week 16 | -0.6 (-2.4 to 1.2) | -0.1 (-1.5 to 1.3) | 0.5 (-1.8 to 2.8) |
| Change at 1-year | -1.5 (-3.9 to 0.9) | -1.2 (-2.8 to 1.4) | 0.3 (-2.9 to 3.5) |
| Fruits (servings/day) | | | |
| Baseline | 1.8 (0.8 to 2.8) | 2.2 (1.2 to 3.1) | - |
| Change at week 16 | 0.1 (-1.2 to 1.4) | 0.8 (-0.8 to 2.4) | 0.7 (-1.4 to 2.8) |
| Change at 1-year | 1.4 (-0.2 to 3.0) | 2.0 (-0.1 to 4.1) | 0.6 (-1.4 to 3.2) |
| Total nuts (servings/day) | | | |
| Baseline | 2.3 (0.6 to 4.0) | 1.8 (0.8 to 2.9) | - |
| Change at week 16 | 0.9 (-1.2 to 3.0) | 2.4 (-0.3 to 4.5) | 1.5 (-1.7 to 4.7) |
| Change at 1-year | 0.5 (-0.8 to 2.3) | 3.3 (-0.6 to 7.2) | 2.8 (-1.4 to 6.9) |
| Legumes (servings/day) | | | |
| Baseline | 3.9 (0.2 to 7.5) | 2.8 (1.9 to 3.7) | - |
| Change at week 16 | -1.1 (-4.7 to 2.6) | 2.3 (-1.6 to 6.2) | 3.4 (-1.9 to 8.7) |
| Change at 1-year | -2.9 (-6.6 to 0.8) | -1.3 (-2.3 to 0.3) | 1.6 (-2.3 to 5.5) |
| Cereals (servings/day) | | | |
| Baseline | 2.1 (1.2 to 2.9) | 1.5 (0.9 to 2.1) | - |
| Change at week 16 | -1.1 (-2.0 to 0.2) | 0.0 (-0.8 to 0.8) | 1.1 (-0.3 to 2.5) |
| Change at 1-year | -1.1 (-2.0 to 0.2) | -0.1 ( -1.0 to 0.8) | 1.0 (-0.4 to 2.4) |
| Olive oil (servings/day) | | | |
| Baseline | 2.4 (1.4 to 3.3) | 2.4 (1.6 to 3.3) | - |
| Change at week 16 | -0.7 (-1.8 to 0.4) | -0.6 (-1.7 to 0.5) | 0.1 (-1.4 to 1.7) |
| Change at 1-year | -1.9 (-2.9 to 0.9) | -1.8 (-2.8 to 0.8) | 0.1 (-2.4 to 2.7) |
| Pastries and cakes (servings/day) | | | |
| Baseline | 9.6 (1.6 to 17.6) | 8.2 (3.0 to 13.4) | - |
| Change at week 16 | -6.8 (-15.0 to 1.4) | -4.4 (-10.1 to 1.3) | 2.4 (-7.6 to 12.4) |
| Change at 1-year | -1.6 (-1.8 to 7.6) | 0.6 (-7.7 to 9.5) | 2.2 (-7.6 to 12.0) |

| | | | |
|---|---|---|---|
| Sugary drinks (servings/day) | | | |
| Baseline | 4.7 (1.6 to 7.9) | 5.0 (0.3 to 9.8) | - |
| Change at week 16 | -1.6 (-10.8 to 7.6) | -4.1 (-7.3 to 0.9) | -2.5 (-12.6 to 7.6) |
| Change at 1-year | -2.1 (-5.5 to 1.3) | -1.5 (-11.4 to 8.4) | 0.6 (-9.9 to 11.1) |
| Alcoholic beverages (servings/day) | | | |
| Baseline | 1.1 (0.5 to 1.7) | 3.0 (0.4 to 5.6) | - |
| Change at week 16 | -0.5 (-1.4 to 0.4) | -0.6 (-4.0 to 2.8) | -0.1 (-3.6 to 3.4) |
| Change at 1-year | -0.5 (-1.4 to 0.4) | -0.6 (-4.0 to 2.8) | -0.1 (-3.6 to 3.4) |

*, $p < 0.05$ time × group interaction; **, $p < 0.01$ time × group interaction.

**Table supplementary S11.** Changes in the food frequency questionnaire after 16-week of supervised exercise intervention and after 1-year post bariatric surgery with intention-to-treat analysis.

| End point | Bariatric surgery + usual care (n=25) | Bariatric surgery + exercise (n=21) | Mean difference between groups |
|---|---|---|---|
| Dairy products (servings/day) | | | |
| Baseline | 2.1 (1.5 to 2.7) | 2.7 (2.0 to 3.4) | - |
| Change at week 16 | -0.4 (-1.4 to 0.6) | 0.0 (-1.2 to 1.2) | 0.4 (-0.1 to 0.8) |
| Change at 1-year | 0.2 (-0.7 to 1.1) | 0.2 (-0.8 to 1.2) | 0.0 (-1.3 to 1.3) |
| Eggs (servings/day) | | | - |
| Baseline | 2.4 (1.7 to 3.0) | 2.5 (1.8 to 3.3) | |
| Change at week 16 | -0.1 (-1.0 to 0.8) | 0.0 (-1.0 to 1.0) | 0.1 (-1.2 to 1.4) |
| Change at 1-year | 11.2 (8.0 to 14.4) | 11.5 (8.5 to 14.5) | 0.3 (-4.1 to 4.7) |
| Meat products (servings/day) | | | |
| Baseline | 17.1 (13.2 to 21.0) | 17.8 (12.9 to 22.6) | - |
| Change at week 16 | -3.5 (-9.0 to 1.5) | -3.8 (-9.7 to 1.9) | 0.3 (-7.5 to 8.1) |
| Change at 1-year | -13.0 (-17.1 to 8.9) | -13.7 (-18.7 to 8.7) | 0.7 (-18.1 to 19.5) |
| Fish or seafood (servings/day) | | | |
| Baseline | 6.2 (4.1 to 8.3) | 9.6 (5.9 to 13.3) | - |
| Change at week 16 | 1.4 (-1.4 to 5.6) | 1.0 (-3.5 to 5.5) | -0.4 (-6.1 to 5.3) |
| Change at 1-year | -4.6 (-6.9 to 2.3) | -7.5 (-11.3 to 3.7) | 2.9 (-5.9 to 11.6) |
| Vegetables (servings/day) | | | |
| Baseline | 4.5 (3.2 to 5.7) | 4.4 (3.3 to 5.4) | - |
| Change at week 16 | -0.6 (-2.3 to 1.1) | -0.1 (-1.5 to 1.3) | 0.5 (-1.7 to 2.7) |
| Change at 1-year | -2.6 (-4.3 to 0.9) | -1.2 (-2.7 to 0.3) | 1.4 (-1.6 to 4.4) |
| Fruits (servings/day) | | | |
| Baseline | 1.8 (0.8 to 2.8) | 3.3 (1.4 to 5.2) | - |
| Change at week 16 | 0.1 (-1.4 to 1.6) | 0.1 (-2.1 to 2.3) | 0.0 (-2.6 to 2.6) |
| Change at 1-year | 1.4 (-0.2 to 3.0) | 1.4 (1.3 to 4.1) | 0.0 (-2.1 to 2.1) |
| Total nuts (servings/day) | | | |
| Baseline | 2.3 (0.6 to 4.0) | 2.1 (1.1 to 3.2) | - |
| Change at week 16 | 0.9 (-1.2 to 3.0) | 2.6 (0.4 to 4.8) | 1.7 (-1.3 to 4.7) |
| Change at 1-year | 0.5 (-0.8 to 2.3) | 3.0 (6.6 to 0.6) | 2.5 (-0.9 to 5.9) |
| Legumes (servings/day) | | | |
| Baseline | 4.5 (0.7 to 8.3) | 6.6 (-1.9 to 15.0) | - |
| Change at week 16 | 1.7 (-5.5 to 2.1) | -1.5 (-10.6 to 7.6) | -3.2 (-13.0 to 6.6) |
| Change at 1-year | -3.6 (-7.4 to 0.2) | -5.1 (-13.6 to 3.4) | 1.5 (-10.8 to 7.8) |
| Cereals (servings/day) | | | |
| Baseline | 2.3 (1.4 to 3.1) | 2.5 (0.6 to 4.4) | - |
| Change at week 16 | -1.4 (-2.3 to 0.5) | -1.0 (-3.0 to 1.0) | 0.4 (-2.0 to 2.8) |
| Change at 1-year | -1.3 (-2.2 to 0.4) | -1.2 (-3.2 to 0.8) | 0.1 (-2.3 to 2.5) |
| Olive oil (servings/day) | | | |
| Baseline | 3.1 (1.7 to 4.5) | 2.8 (1.6 to 4.1) | - |
| Change at week 16 | -1.4 (-3.0 to 0.2) | -1.0 (-2.4 to 0.4) | 0.4 (-1.7 to 2.5) |
| Change at 1-year | -2.6 (-4.1 to 1.1) | -2.1 (-3.5 to 0.8) | 0.5 (-2.9 to 3.9) |
| Pastries and cakes (servings/day) | | | |
| Baseline | 9.2 (1.5 to 16.9) | 8.0 (3.6 to 12.4) | - |
| Change at week 16 | -6.4 (-14.4 to 1.6) | -4.1 (-10.4 to 2.2) | 2.3 (-7.9 to 12.5) |
| Change at 1-year | -1.2 (-10.2 to 7.8) | 0.6 (6.6 to 8.4) | 1.8 (-7.2 to 10.8) |

| | | | |
|---|---|---|---|
| Sugary drinks (servings/day) | | | |
| Baseline | 4.8 (1.7 to 7.8) | 5.7 (1.3 to 10.0) | - |
| Change at week 16 | -2.2 (-5.6 to 1.2) | -2.2 (-7.3 to 2.9) | 0.0 (-6.1 to 6.1) |
| Change at 1-year | -4.2 (-7.3 to 1.1) | -3.6 (-8.4 to 1.2) | 0.6 (-5.8 to 6.9) |
| Alcoholic beverages (servings/day) | | | |
| Baseline | 1.0 (0.5 to 1.6) | 3.0 (0.7 to 5.2) | - |
| Change at week 16 | -0.4 (-1.2 to 0.5) | -0.9 (-3.9 to 2.1) | -0.5 (-3.6 to 2.6) |
| Change at 1-year | -0.4 (-1.2 to 0.5) | -0.9 (-3.9 to 2.1) | -0.5 (-3.6 to 2.6) |

*, $p < 0.05$ time × group interaction; **, $p < 0.01$ time × group interaction.

**Table S12.** Forward stepwise regression models assessing the independent association of the predictors (changes in body weight, body composition, physical fitness, cardiometabolic and inflammatory markers, and metabolomic profile) with changes in SHBG, oocyte count and UtA-PI at 1-year post bariatric surgery for usual care group.

| | **β** | **B** | **95%CI** | ***p*** | **$R^2$** | **$R^2$ change** | **p (model)** |
|---|---|---|---|---|---|---|---|
| ***SHBG*** | | | | | | | |
| **Step 1** | | | | | 0.46 | - | 0.007 |
| 3-HB | 0.68 | 1.41 | 0.46, 2.37 | 0.007 | | | |
| **Step 2** | | | | | | | |
| 3-HB | 0.67 | 1.39 | 0.76, 2.02 | <0.001 | 0.79 | 0.33 | 0.002 |
| Leptin | -0.57 | -0.38 | -0.58, -0.18 | 0.002 | | | |
| **Step 3** | | | | | 0.90 | 0.11 | 0.007 |
| 3-HB | 0.66 | 1.37 | 0.92, 1.83 | <0.001 | | | |
| Leptin | -0.62 | -0.41 | -0.56, -0.27 | <0.001 | | | |
| Ast | -0.34 | -0.35 | -0.57, -0.12 | 0.007 | | | |
| ***Oocyte count*** | | | | | | | |
| **Step 1** | | | | | 0.45 | - | 0.016 |
| Bruce test | -0.67 | -1.56 | -2.76, -0.35 | 0.016 | | | |
| **Step 2** | | | | | 0.74 | 0.28 | 0.012 |
| Bruce test | -0.69 | -1.56 | -2.49, -0.71 | 0.003 | | | |
| Creatine | 0.53 | 2.87 | 0.79, 4.96 | 0.012 | | | |
| **Step 3** | | | | | 0.86 | 0.12 | 0.033 |
| Bruce test | -0.82 | -1.89 | -2.65, -1.13 | <0.001 | | | |
| Creatine | 0.56 | 3.04 | 1.37, 4.71 | 0.003 | | | |
| LDL | 0.37 | 1.11 | 0.12, 2.10 | 0.033 | | | |
| ***UtA-PI*** | | | | | | | |
| **Step 1** | | | | | 0.42 | - | 0.009 |
| Aspartate | -0.65 | -0.89 | -1.5, -0.26 | 0.009 | | | |
| **Step 2** | | | | | 0.62 | 0.20 | 0.029 |
| Aspartate | -0.61 | -0.84 | -1.37, -0.30 | 0.005 | | | |
| PWV | -0.44 | -2.20 | -4.14, -0.26 | 0.029 | | | |
| **Step 3** | | | | | 0.88 | 0.26 | <0.001 |
| Aspartate | -0.26 | -0.36 | -0.75, 0.03 | 0.066 | | | |
| PWV | -0.66 | -3.27 | -4.53, -2.01 | <0.001 | | | |
| TAG | -0.65 | -0.64 | -0.93, -0.35 | <0.001 | | | |
| **Step 4** | | | | | 0.93 | 0.05 | 0.025 |
| Aspartate | -0.31 | -0.42 | -0.74, -0.10 | 0.015 | | | |
| PWV | -0.75 | -3.73 | -4.83, -2.63 | <0.001 | | | |
| TAG | -0.57 | -0.56 | -0.81, -0.31 | <0.001 | | | |
| Leptin | -0.26 | -0.11 | -0.21, -0.02 | 0.025 | | | |
| **Step 5** | | | | | 0.97 | 0.04 | 0.012 |
| Aspartate | -0.48 | -0.66 | -0.96, -0.37 | <0.001 | | | |
| PWV | -0.80 | -3.95 | -4.78, -3.12 | <0.001 | | | |
| TAG | -0.60 | -0.59 | -0.78, -0.41 | <0.001 | | | |
| Leptin | -0.29 | -0.13 | -0.20, -0.06 | 0.003 | | | |
| GlyOH | 0.28 | 0.87 | 0.24, 1.50 | 0.012 | | | |
| **Step 6** | | | | | 0.99 | 0.02 | 0.010 |
| Aspartate | -0.52 | -0.72 | -0.93, -0.51 | <0.001 | | | |
| PWV | -0.83 | -4.13 | -4.72, -3.54 | <0.001 | | | |
| TAG | -0.61 | -0.61 | -0.74, -0.48 | <0.001 | | | |
| Leptin | -0.31 | -0.13 | -0.19, -0.08 | <0.001 | | | |
| GlyOH | 0.26 | 0.79 | 0.35, 1.23 | 0.003 | | | |

| | | | | | | | |
|---|---|---|---|---|---|---|---|
| Chair Stand | -0.16 | 0.47 | -0.79, -0.15 | 0.010 | | | |
| **Step 7** | | | | | 0.99 | 0.01 | 0.013 |
| Aspartate | -0.49 | -0.67 | -0.82, -0.53 | <0.001 | | | |
| PWV | -0.81 | -4.01 | -4.43, -3.60 | <0.001 | | | |
| TAG | -0.66 | -0.66 | -0.75, -0.56 | <0.001 | | | |
| Leptin | -0.28 | -0.12 | -0.16, -0.09 | <0.001 | | | |
| GlyOH | 0.30 | 0.94 | 0.62, 1.26 | <0.001 | | | |
| Chair Stand | -0.14 | -0.41 | -0.63, -0.18 | 0.003 | | | |
| Cortisol | 0.12 | 0.07 | 0.02, 0.12 | 0.013 | | | |

B, standardized regression coefficient; B, unstandardized regression coefficient; $R^2$, adjusted coefficient of determination, expressing the percent variability of the dependent variable explained by the model; $R^2$ change, additional percent variability explained by the model due to the inclusion of the new predictor; 3-HB, 3-hydroxybutyrate; Ast, aspartate transaminase; LDL, low-density lipoprotein; UtA-PI, uterine artery mean pulsatility index; PWV, pulse wave velocity; TAG, triglycerides; GlyOH, glycolic acid.

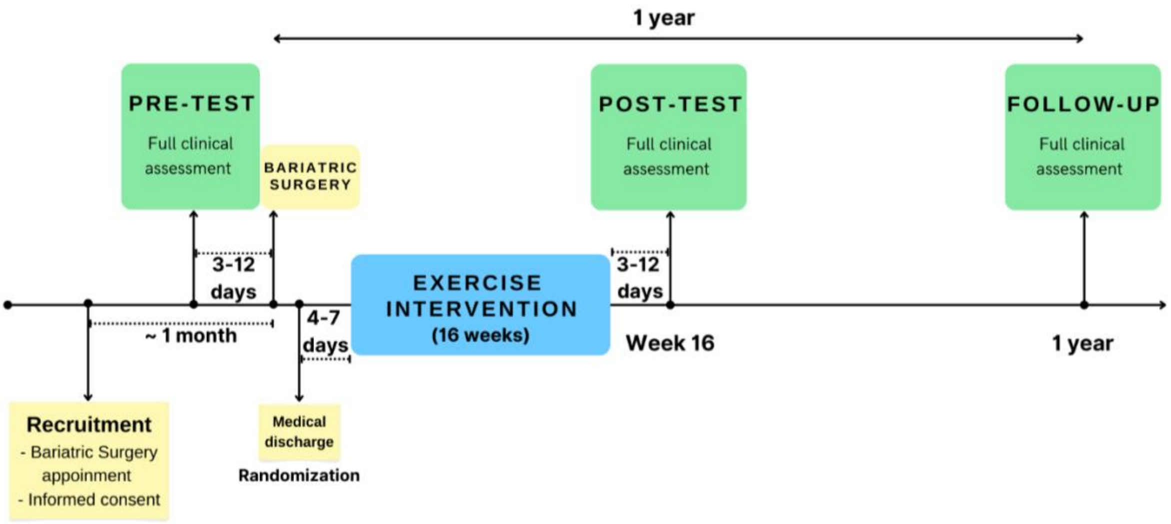


**Figure S1.** Graphical representation of the timeline of main events during the EMOVAR randomized trial.

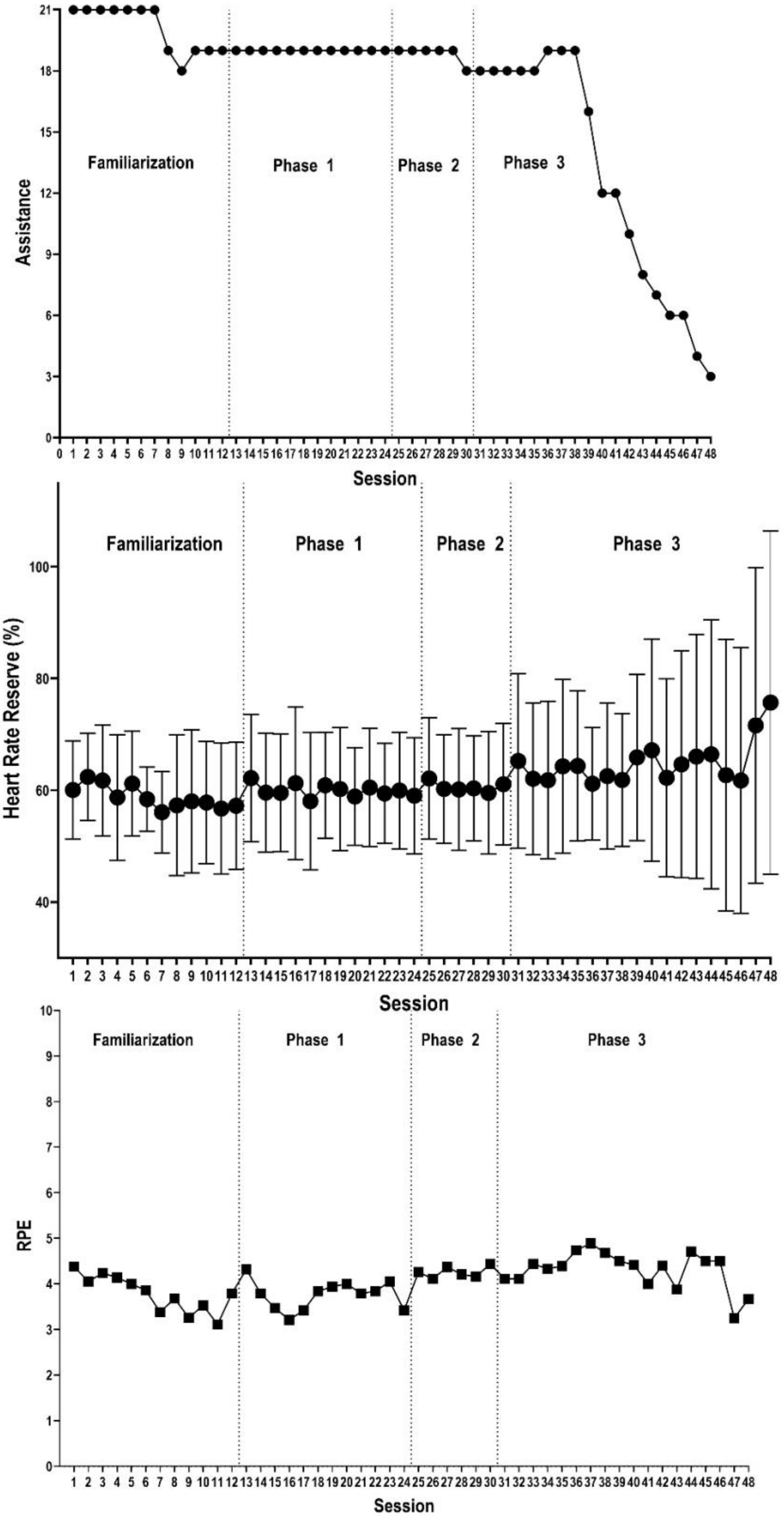


**Figure S2**. Summary of participants assistance, objective (i.e., heart rate) exercise intensity and session rating of perceived exertion (RPE) during each session of the exercise intervention.

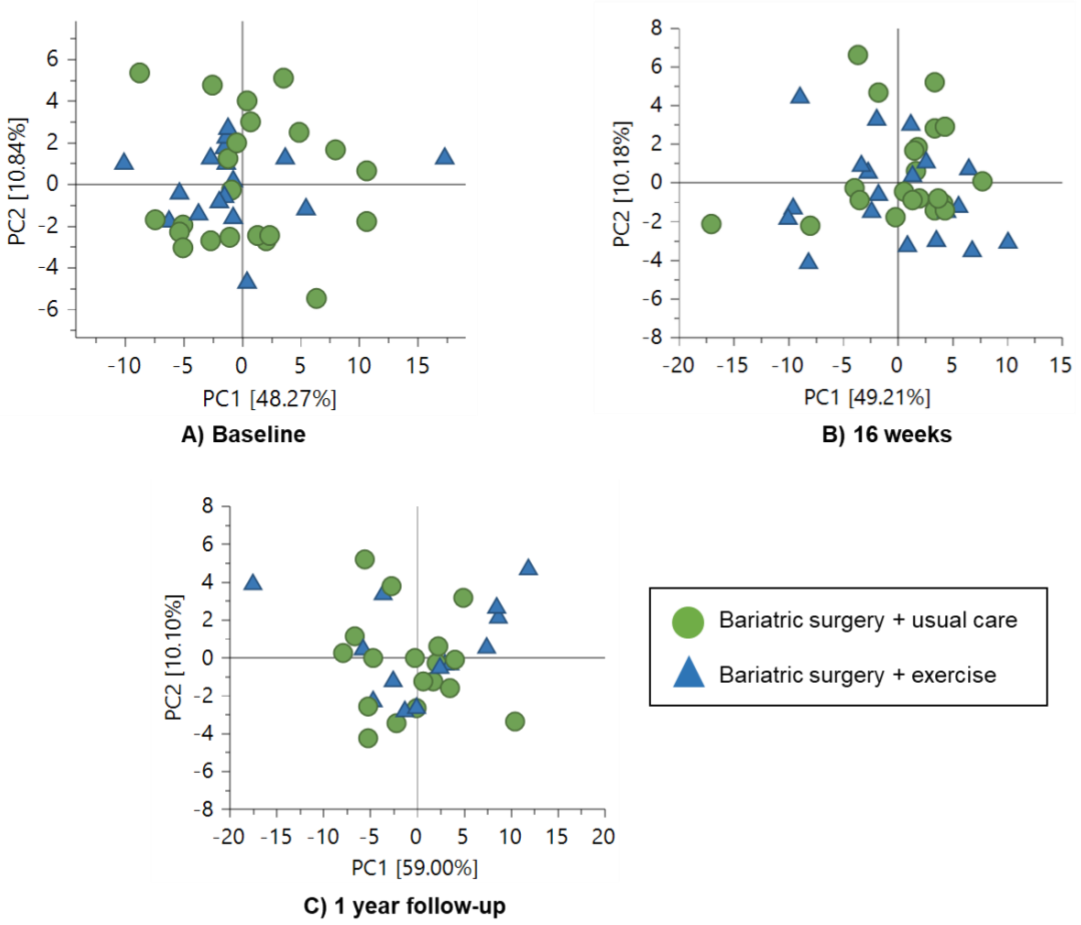


**Figure S3.** Principal Component Analysis (PCA) scores plots comparing NMR spectra of both groups (BS plus usual care *vs.* BS combined with exercise) across intervention end points: A) before surgery, B) week 16, and C) 1-year post bariatric surgery.

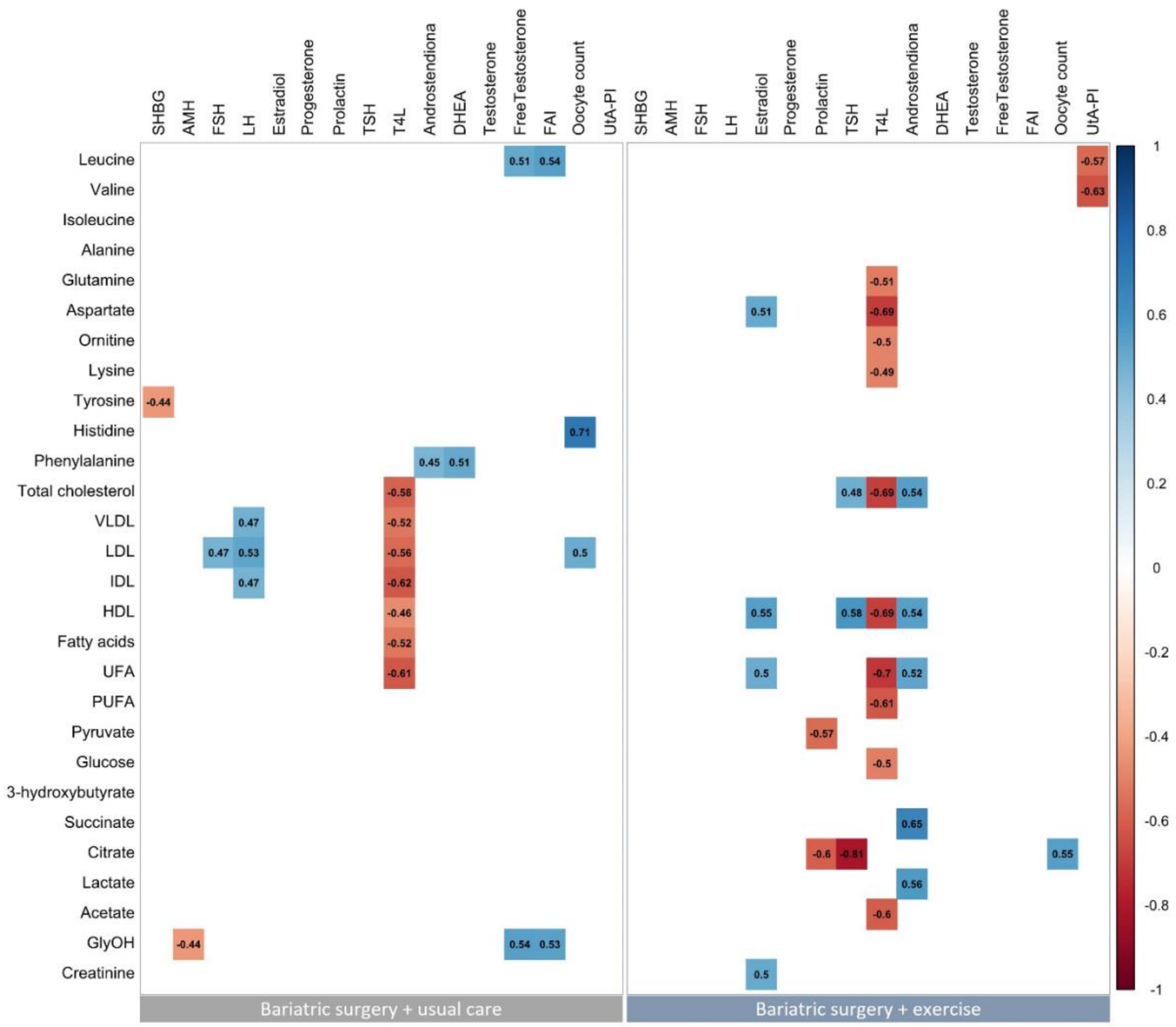


**Figure S4.** Pearson correlation analyses between changes in metabolites (deduced from NMR) with changes in ovarian function after the 16-week exercise intervention. Every box represents a significant correlation coefficient (all $p < 0.05$), whereas empty boxes represent no significant correlations.
